# Hospitalisation and mortality impact of shielding during 2020 in England: a transmission modelling evaluation using the OpenSAFELY platform

**DOI:** 10.64898/2025.12.12.25342168

**Authors:** Johnny A. N. Filipe, Edwin van Leeuwen, Alasdair Henderson, Nicholas G Davies, Christopher Jarvis, Helen J Curtis, Koen B Pouwels, W John Edmunds, Brian MacKenna, Seb Bacon, Amir Mehrkar, Ben Goldacre, Laurie Tomlinson, Rosalind M Eggo

## Abstract

**Background:** During the early phase of the Covid-19 pandemic in England, people with pre-existing conditions that put them at severe clinical risk if infected were advised to drastically reduce face-to-face contacts in a policy known as “shielding”. The impact of this policy in preventing COVID-19 hospitalisations and deaths has not been evaluated at the national level using transmission-dynamic modelling.

**Methods:** With the approval of NHS England, we present a retrospective cohort evaluation of the shielding policy, drawing data from electronic health records (EHRs) for 24 million patients in England accessed through the OpenSAFELY platform. The study is from 1 January to 1 December 2020, prior to vaccination and new SARS-CoV-2 variants. We used a dynamic transmission model of SARS-CoV-2 transmission, infection, and hospitalisation, stratified by age and shielding status for the general population (excluding care homes). We estimated transmission rates in the shielding and non-shielding groups using data from the CoMix social contact survey, and fitted the model to hospitalisations and deaths in and outside hospital.

**Findings:** We found that the risk of hospitalisation was higher for shielding people in all age groups and increased with age. The hospital fatality ratio was similar between shielding and non-shielding people from January to June 2020 and greater in shielding people from July 2020 onward. By comparing the observed epidemic to a counterfactual scenario without shielding, we projected that between 7800 and 10,600 hospitalisations and 2300 to 3500 deaths due to COVID-19 were directly averted by the policy, corresponding to reductions of 25% (24, 28%) and 23% (21, 25%), respectively, in the shielding population in England up to 1 December 2020. Including also the indirect effect in the non-shielding population, we projected between 14,700, and 21,800 hospitalisations and 3700 and 5500 deaths due to COVID-19 were averted by the policy in the total population, each corresponding to reductions of 13% (11, 16%).

**Interpretation:** Based on the data and assumptions in this study, the shielding policy reduced pressure on the NHS and severe illness and mortality in clinically-extremely vulnerable shielding patients in England up to 1 December 2020, and, through indirectly-reduced exposure, also in the non-shielding population. Similar policies for other infections could have a comparable public health impact in reducing both mortality and pressure on public health services.

**Funding:** Medical Research Foundation, Medical Research Council, National Institute for Health and Care Research, NHS England, The Wellcome Trust.

## Introduction

During the first part of the 2020 COVID-19 pandemic in the UK, people identified as clinically extremely vulnerable (CEV) to severe illness from COVID-19 were advised to self-isolate by staying at home and avoiding all non-essential face-to-face contact^1,2^. This UK shielding programme started on 22 March 2020 and provided CEV people with public health guidance and support in accessing food, medicine and basic care. The policy aimed to prevent infection by SARS-CoV-2 and thereby prevent severe illness and death in these at-risk groups, and reduce pressure on the NHS [1]. In England, the Shielded Patient List (SPL) contained over two million CEV patients and remained at around this level throughout 2020^3,4^.

Evaluations of the impact of shielding in the first phase of the pandemic compared COVID-19 outcomes between comparatively small shielding and non-shielding cohorts in England^5^, Wales^6^ and Scotland^7^, and a larger study in Wales^8^. These observational studies are limited as they cannot estimate how the risk of infection differs in shielding and non-shielding groups and therefore cannot control for the specific risk of severe outcomes separately from the specific risk of infection in each cohort. Here, we aimed to estimate the population-level impact of the policy in a large CEV cohort by jointly estimating the dynamic risks of infection and of a severe outcome upon infection. The first depends on the community prevalence of SARS-CoV-2 infection and on the level of exposure (i.e. “social contacts”), both of which varied rapidly and widely throughout the 2020 pandemic.

Electronic Health Records (EHRs) offer comprehensive patient-level clinical data without common surveillance data limitations, such as under-reporting and temporal or geographic aggregation. We used EHRs from 24 million NHS patients accessed via OpenSAFELY^9^, combined with data from the CoMix social contact survey^10,11^, dynamic transmission modelling, and Bayesian inference, to evaluate the impact of the shielding policy in England. We estimated the impact by comparing the CEV cohort with a counterfactual scenario where the CEV group had the same contacts as the non-shielding group. Our study period is 27 January to 01 December 2020, i.e. from the start of the pandemic to the early stages of its second wave.

## Methods

We estimated the dynamic risk of infection and the risk of a severe outcome following infection by fitting a mathematical model of SARS-CoV-2 transmission to age– and shielding-stratified hospitalisation events and mortality drawn from EHRs. We used social contact rates measured over the study period to estimate the time-varying risk of SARS-CoV-2 infection by age and shielding status. To infer the effect of the shielding policy we simulated a counterfactual scenario where those in the CEV group had the same social contact rates as the general population.

### Electronic Health Records data

All data were linked, stored and analysed securely using the NHS England OpenSAFELY COVID-19 service [8]. Data include pseudonymised data such as coded diagnoses, medications and physiological parameters. No free text data are included. Pseudonymised primary-care patient records were linked to hospital admissions, mortality, care home residence, and information on a patient’s shielding status (published by PRIMIS (Primary Care Information Services, University of Nottingham)). All analysis code is shared openly for review and re-use under MIT open license (https://github.com/opensafely/shielding-evaluation). Detailed pseudonymised patient data is potentially re-identifiable and therefore cannot be shared.

We used EHR records from patients registered at general practices (GP) that use SystmOne software from provider TPP^9^. We extracted data from 1 January–1 December 2020 (48 weeks) but used data starting on 27 January 2020 (45 weeks) to match the modelled epidemic start date. We included patients aged 0-100 years, alive at the start of the study period, registered at a GP for at least 90 days, and who had a single registration by the end of the study, giving 24.02 million patients (excluding 233,865 patients with missing demographic data). We further excluded 51,810 (0.22%) patients residing in care homes because social contacts likely differed in these facilities. The final study population had 23.97 million patients, representing 42.5% of the population of England in 2020.

From the extracted data we derived the weekly incidence of COVID-19-associated severe outcomes: hospital admissions, deaths in hospital, and deaths outside hospital, each stratified into 9 age groups (Table S2) and shielding status. Statistics on the severe outcome data (Table S2) and details of the extraction, including information on codelists used to identify COVID-19 outcomes, are in Supplementary Section 1. To fit the model, we scaled up the number of events in each week and age group from the 42.5% sample of patients in TPP practices^13^, to 100% of the England population excluding those in care homes. All figures show data for England, and scaling occurred after non-disclosure control rounding had been applied. The model was fitted to data that had non-disclosure controls applied (Supplementary Section 8).

### Shielding population

In England, 2.25 million NHS patients were identified in March 2020 as CEV due to pre-existing health conditions. A code was added to the GP record of these patients^3^. There was some discretion for GPs to add patients, but the majority were added centrally. While people could be added to the CEV list due to onset of an eligible condition or prescription, and the list of conditions indicating CEV was later also reviewed, the online SPL dashboard showed that the number of CEV-identified patients remained stable from March 2020 until the end of our study period^14–16^, at around 4% of the population in England. The percentage shielding varied by age from 0.7% in 0-4 year olds, rising to 13.7% in the 70+ age group (Supplementary Section 2). Based on the stable net number of CEV-identified patients on the SPL, we assumed that patients did not change shielding status during the study period.

### Transmission model

We developed a deterministic compartmental model of SARS-CoV-2 transmission among the 56.43 million residents in England (excluding those in care homes) in 2020^12^ (Figure 1a) stratified by age group (*i*=1…9) and shielding status (*s*=yes/no). The model extends existing work (similar to^17,18^) by including shielding stratification, time-varying social contacts drawn from survey data, and a second pathway to mortality in the community that bypasses hospitalisation.

**Figure 1.**
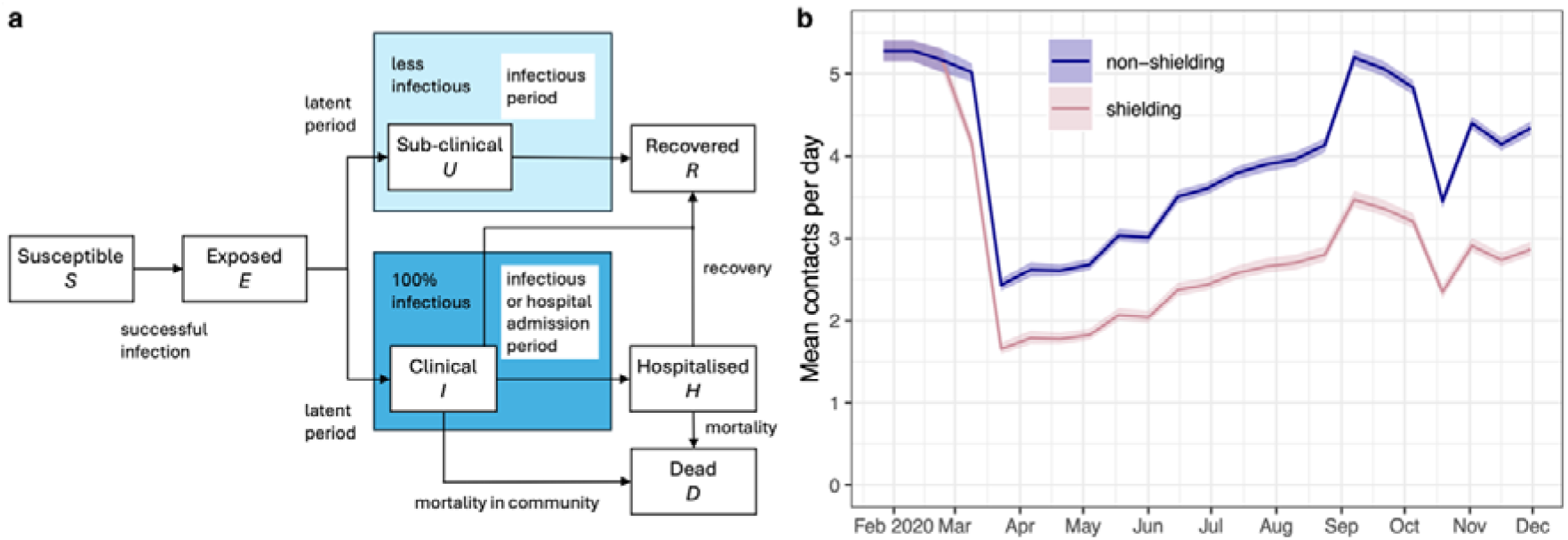
Transmission model details. a) States and transitions of the SARS-CoV-2 transmission model. People could be in the Susceptible, Exposed, Sub-clinically infected, Clinically-infected, Recovered (and immune), Hospitalised, or Dead states. Each state compartment is stratified into 9 age bands and yes/no shielding groups. The epidemic was simulated starting on 27 January 2020 with a number of infected individuals that was estimated through model fitting. All individuals were initially susceptible (*S*) with no pre-existing immunity to SARS-CoV-2 infection, and became exposed (*E*) upon effective contact with an infectious individual. b) Mean daily number of contacts in the non-shielding and shielding populations in England from 27 January to 01 December 2020 derived from the CoMix social contact survey (see Methods). The shielding policy started on 22 March 2020. We show the mean across age groups and 95% credible interval (based on the posterior sample of the fitted regression model) here, but in the model, age-specific contact patterns were used.

All individuals were susceptible at the start of the study, and when infected could exhibit subclinical (*U*) or clinical infections (*I*). Sub-clinical infections are those with very few or no symptoms and thus are assumed to be unascertained, whereas those displaying clinical infections may (or may not) seek care. We estimated the probability of subclinical vs clinical infection and it differed by age and shielding status (Supplementary Section 8 and 12). Clinically-infected individuals either became hospitalised (*H*) after an average hospital-admission period, or died in the community (*D*) after an average outside-hospital mortality period, or recovered (*R*). Hospitalised individuals either progressed to recovery or died.

We estimated the infectiousness of sub-clinical infections relative to clinical infections and assumed that all individuals recovered^18,19^. We assumed no waning of immunity to re-infection during the study period. Further details of the model are given in Supplementary Section 3.

### Model parameters

We assumed that the susceptibility to infection depended on age^17^, and that the clinical probabilities of showing symptoms (clinical infection), hospitalisation, and mortality (in hospital and community separately) differed by age and shielding groups and could be estimated through model fit (except the hospital fatality ratio which was calculated directly from EHR data). These include two widely used epidemiological indicators: the hospitalisation fatality risk (HFR) and the infection hospitalisation risk (IHR). We started epidemics on 27 January 2020 (^17,19^) by seeding infected individuals among all age groups and estimated the number during inference. The basic reproductive number (*R*_0_) was calculated from the next generation matrix. Further details are given in Supplementary Section 14.

### Social contact survey data

CoMix was an online social contact survey of representative cross-sectional samples of the UK population, with responses from all participants reported every 2 weeks from 24 March 2020, the day after the start of the first national lockdown^10,11^. Children were included in the survey starting two months later. Participants responded to a questionnaire about the frequency and nature of their daily face-to-face contacts on the day and in the previous week, about their age and approximate age of their contacts, and about their circumstances and attitudes. We used these data to parameterise a time-varying contact matrix in the transmission model (Figure 1), stratified by age and shielding groups. We used questions about the participant’s perception of risk to identify which survey participants were potentially shielding and CEV patients and thus estimated their social contacts separately from those of the non-shielding population (Figure 1b). We extracted biweekly estimates of mean daily contacts of residents in England stratified by the age groups of the participant and of the contact.

To extrapolate to the start of our study period (27 January 2020) and to tackle the delayed start of data collection for children we used Bayesian regression with covariates that tracked activity (in schools, workplaces and recreation places) and temperature over the study period (Supplementary Section 6). As the shielding policy officially started on 22 March 2020^1,2^, we assumed that CEV-identified people were aware of risk and started to shield gradually from three weeks earlier. The overall level of social contacts in the general and shielding populations dropped dramatically during the first and second national lockdowns, but the reduction was greater in the shielding population (Figure 1b). Further details are given in Supplementary Section 6.

### Model fitting

We used Bayesian evidence synthesis^20^ to fit the transmission model to weekly incidences of severe outcomes and infer the risks of hospitalisation and death in and outside hospital. We fitted the model to 54 weekly time series of COVID-19 outcomes drawn from EHR data (3 outcomes, 9 age groups and 2 shielding strata). We sampled the parameter joint posterior distribution using a negative binomial likelihood function and the Differential Evolution^21,22^ implementation of Metropolis-Hastings Markov chain Monte Carlo (MCMC)^23^, using the R package BayesianTools^24^. We ran 3 chains for 400,000 iterations, where 50% were used as burn in. We report posterior medians and 95% credible intervals (95% CrI) for the parameters, and medians and 95% prediction intervals (95% PI) of posterior predictive distributions for comparison with observed data. The latter distributions were obtained via simulation of the observations upon sampling from the joint posterior samples. Further details of the fitting approach, MCMC diagnostics, and parameter posterior outcomes are given in Supplementary Section 10.

### COVID-19 positivity survey data

During the lockdown period, the UK’s Office for National Statistics Covid Infection Survey (ONS-CIS) sent swab tests to a representative sample of the general population^25^. These data are available publicly and give an estimate of infection prevalence in the general population^26^. As a validation of our transmission model, we used publicly available swab-positivity data for England to check alignment with model outputs. We did not fit to ONS-CIS prevalence. Further details on the survey are given in Supplementary Section 9.

### Evaluation of shielding

To estimate the impact of the shielding policy, we simulated the weekly incidence of severe COVID-19 outcomes in England in a scenario where CEV-identified people had contact patterns the same as age-matched non-shielding people. We simulated the epidemic for each sample from the joint posterior distribution of the fitted model (Tables S5 and S6), using the same initial condition and the same fixed parameters. For each posterior parameter point, we calculated the numbers of hospitalisations and deaths (inside and outside hospital) and their difference with respect to the fitted model. We separately considered severe outcomes averted in the shielding and the non-shielding population (Table 3).

### Sensitivity analysis

We tested how sensitive our findings were to the assumption that social contacts were Poisson distributed (Supplementary Section S6) compared with an assumption of a Negative Binomial distribution. Additionally, the CoMix data are the best available information on the social contact patterns of people at risk of severe outcomes of infection, however, to determine the sensitivity of our conclusions, we tested a model where contacts were reduced by a further 10 or 20% beyond that reported in CoMix.

### Ethics

This study was approved by the Health Research Authority (Research Ethics Committee reference 20/LO/0651) and by the London School of Hygiene & Tropical Medicine Ethics Board (reference 29468).

### Patient and public involvement

OpenSAFELY has involved patients and the public through multiple routes: there is a public website that provides a detailed description of the platform (https://opensafely.org); the core OpenSAFELY team have participated in two citizen juries exploring public trust in OpenSAFELY, and have co-developed an explainer video (https://www.opensafely.org/about/);. There is patient representation through people who are experts by experience on the OpenSAFELY Oversight Board, and there is an ongoing partnership with Understanding Patient Data to produce lay explainers on the importance of large datasets for research. OpenSAFELY have presented at online public engagement events to key communities. We are committed to ensuring patient voices are heard in research.

## Results

### Characteristics of the cohort

The age distribution of the cohort was similar to ONS population estimates for mid 2020^12^ (Table S1). The cohort was not evenly distributed across all NHS England regions. However, previous studies have reported that patients registered in TPP-served GPs are representative of the English population in key demographic characteristics^13^. We identified 1,006,960 patients with a CEV code (4.2%) in the study population, close to the 4% of CEV patients in the English population. The proportion of people shielding increased with age, rising from 0.7% in 0-4 year olds to 13.7% in the 70+ age group (Table S1). Among 42% of the England population, we identified 50,770 hospitalisations, of which 48% were in people over 70 years old, and 16,940 deaths, 4,250 of which were outside hospital (Table S2). Scaling to the entire population of England, this would give 119,400 hospitalisations, 30,400 deaths in hospital and 10,400 deaths outside hospital (in non-care home residents).

### Fit to weekly incidence of COVID-19 outcomes

We fitted the model to data for 9 age groups but, for simplicity, display the model estimates aggregated across all age groups, separated by shielding status (Figure 2). The cumulative model estimates for the entire population of England were 115,000 (median and 95% PI 53,400-213,400) hospitalisations, 21,100 (5300-51,200) deaths in hospital, and 5800 (200-30,200) deaths outside hospital. The majority of hospitalisations and deaths were in people over 60 in the study period, and the model fits both groups well (Figure 3). As expected, the data show considerably more hospitalisations and especially deaths in the older adult patients (Table S2). The model 95% PI contain the data and the median posterior curves follow the data patterns reasonably closely. Age-specific fits are shown in Supplementary Section 11.

**Figure 2.**
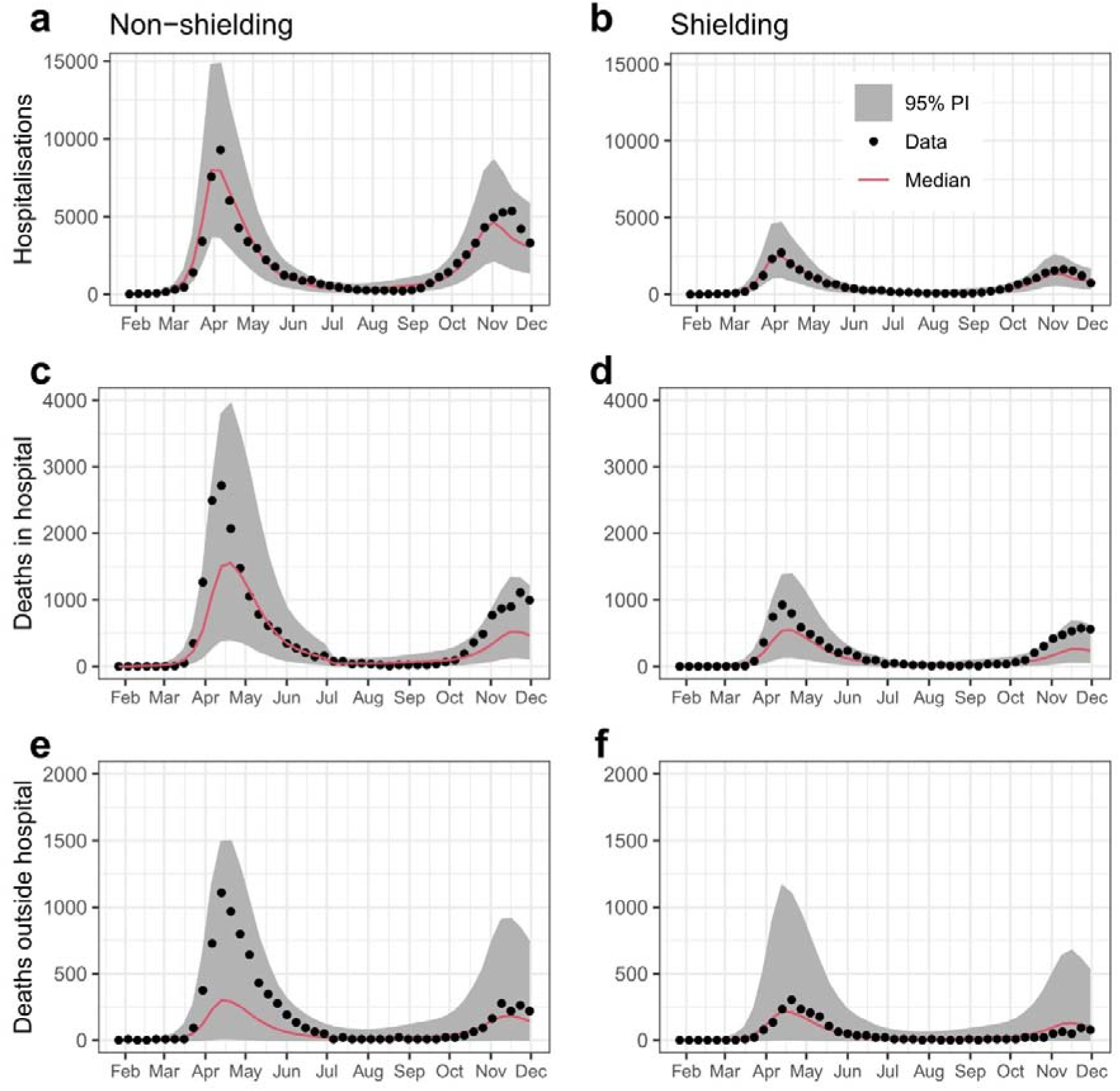
Weekly incidence of COVID-19 hospitalisations (a, b), deaths in hospital (c, d), and deaths outside hospital (e, f) in England in the non-shielding and shielding populations. Red lines and grey shading shows posterior predictive distributions (median and 95% PI) of the fitted transmission model, black dots are data drawn from OpenSAFELY aggregated across all ages. Note that the model was fitted to age-stratified data.

**Figure 3:**
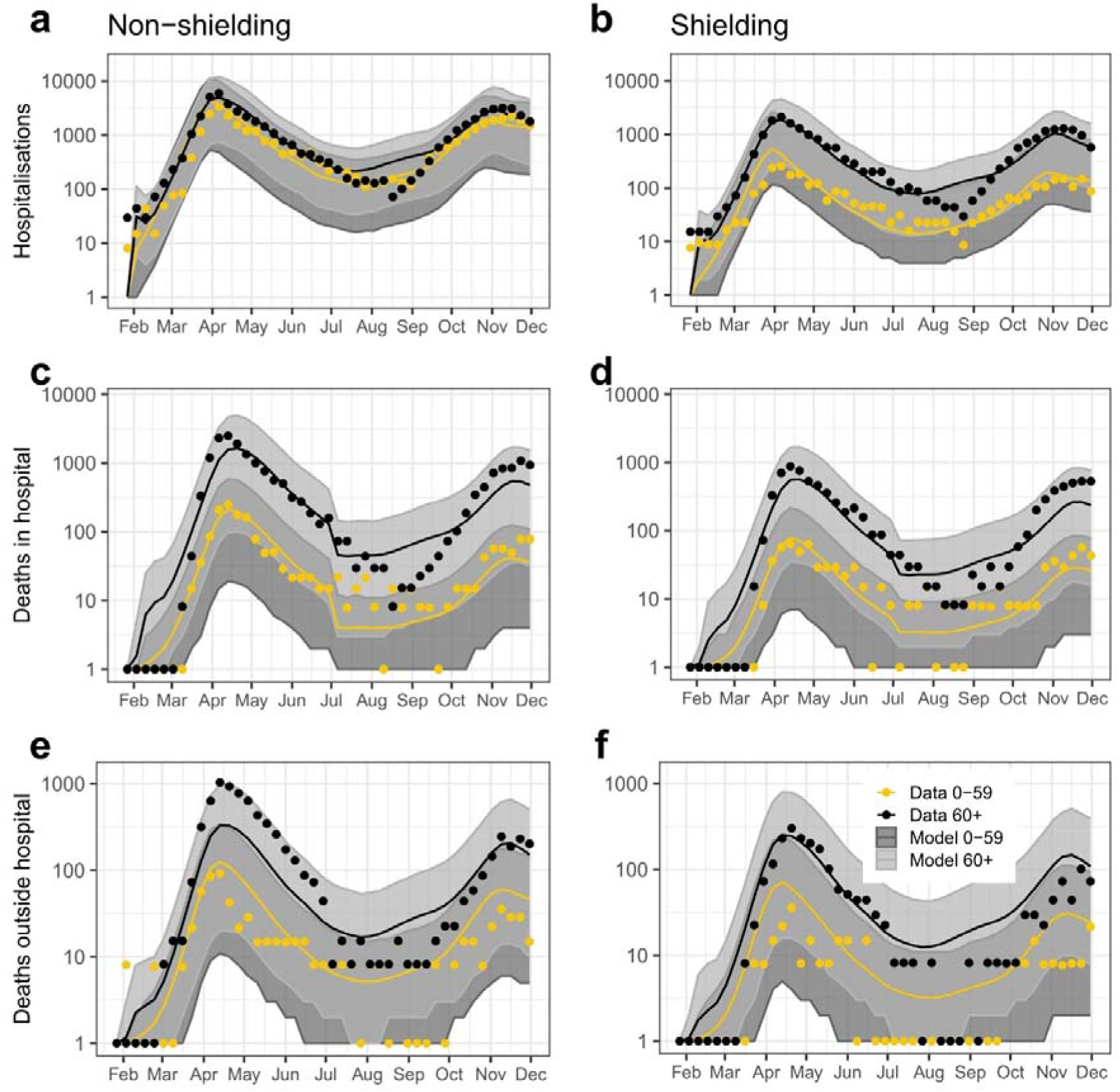
Weekly incidence of COVID-19 hospitalisations (a, b), deaths in hospital (c, d), and deaths outside hospital (e, f) in England for ages 0-59 and 60+ years of age in the non-shielding and shielding populations. The model and data are shown on a log-10 scale. Posterior predictive distribution of the fitted transmission model (median (line) and 95% prediction interval (grey area)) and data from OpenSAFELY (dots). Mid grey indicates overlap between 0-59 and 60+ prediction intervals. For plotting on log10 scale, a lower cut-off of 1 was applied. Note that the model was fitted to 9 age groups but is shown aggregated to 2 here.

### Clinical responses to COVID-19 by age and shielding status

The parameters of the fitted model show that the risk of developing clinical illness following infection, of being subsequently hospitalised, and the combination of these two risks (or IHR) were higher for shielding people in all age groups and increased with age for shielding and non-shielding people (Figure 4a). The inferred risk of dying outside hospital upon developing clinical infection (Figure 4b) was considerably higher for shielding people; this group may have included ill people that were too frail to be hospitalised. These results are consistent with the expectations that shielding people were at greater clinical risk from COVID-19 and that these risks increased with age. Because we separately calculated the risk of death after admission to hospital (HFR) before and after June 2020, we found that the HFR was similar between shielding and non-shielding people from January to June 2020 and greater in shielding people from July to December 2020 (Figure 4c).

**Figure 4:**
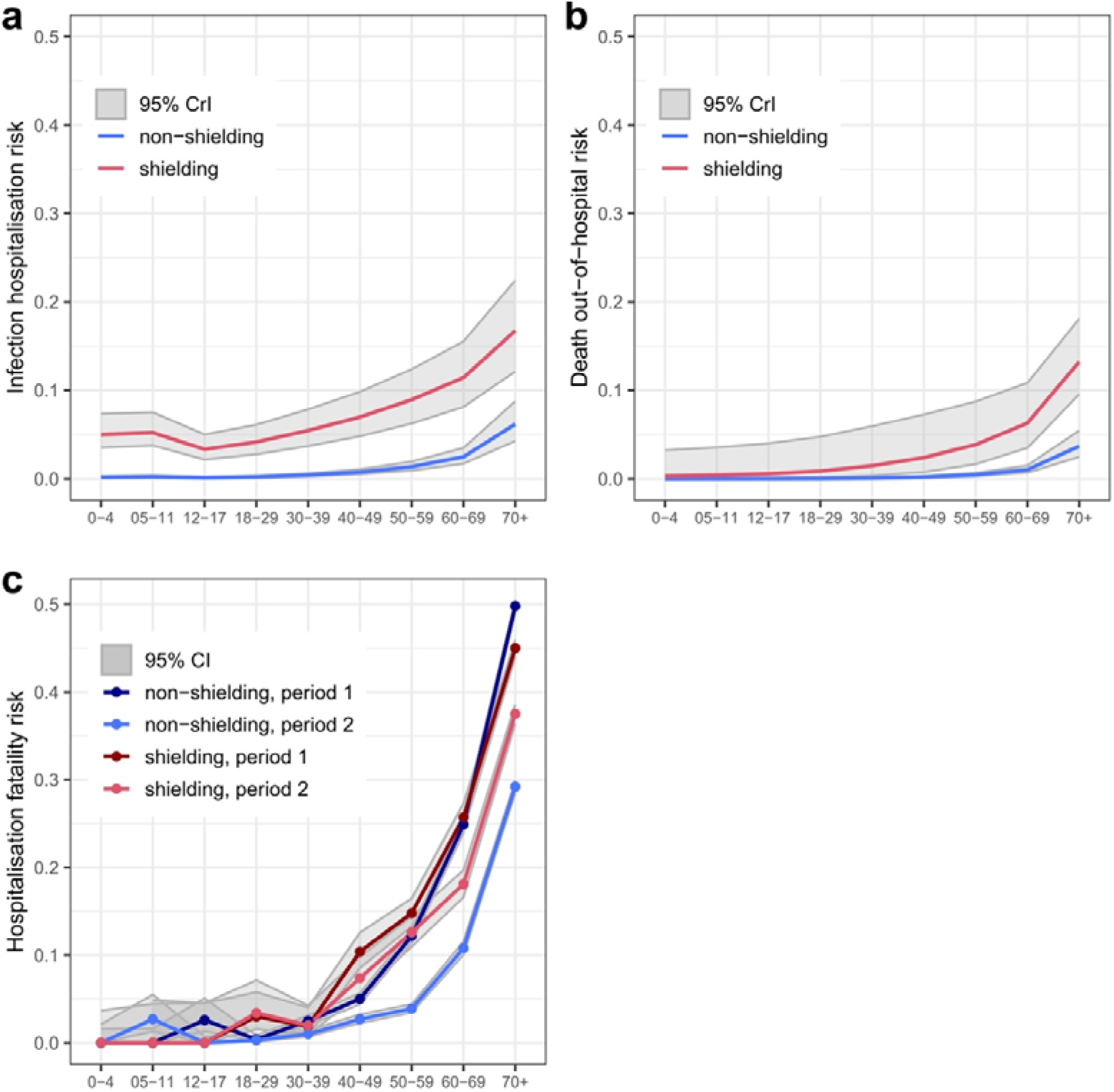
Age-specific estimates of key epidemiological parameters in shielding and non-shielding groups. a) Infection hospitalisation risk is the risk of death following infection (clinical or non-clinical), b) risk of death out-of-hospital in those infected, c) hospitalisation fatality risk. a and b were inferred during model fitting, and c was calculated directly from EHR data. Posterior medians and 95% credible intervals are shown for shielding (red) and non-shielding (blue) populations, where the HFR was calculated for two time periods in c.

### Predicted versus observed positivity

As an external validation of the fit of the model, we compared the model prediction of SARS-CoV-2 positivity in England with the ONS Coronavirus Infection Survey and found good agreement (Figure 5).

**Figure 5:**
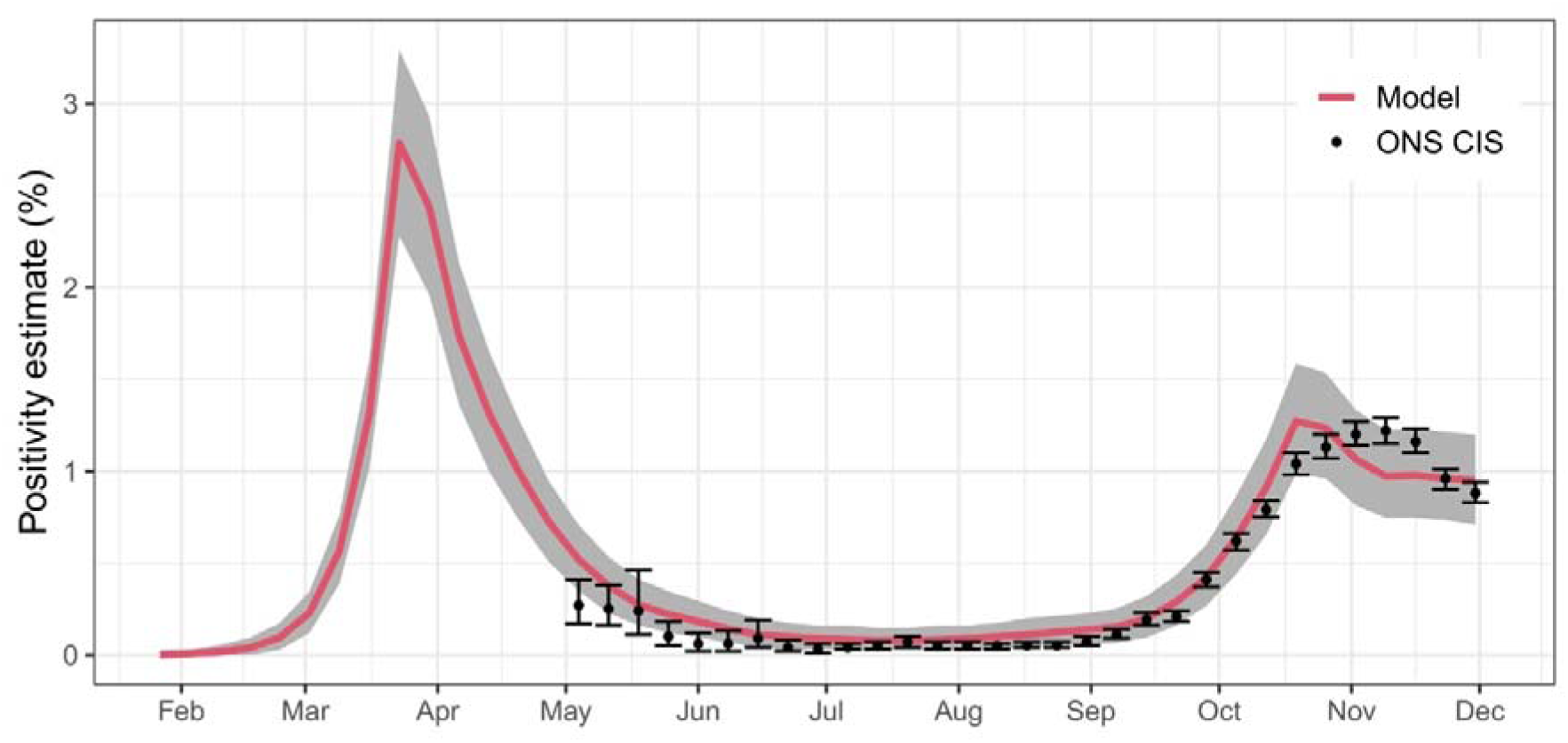
Weekly PCR-positivity of COVID-19 in England from 27 January to 01 December 2020. The non-shielding and shielding populations are included in the estimates. Model weekly prediction of virus positivity (median and 95% prediction interval, 95% PI) and observed percentage from the ONS Coronavirus Infection Survey of people (symptomatic or non-symptomatic) with a positive PCR test starting on 03 May 2020 (average and 95% confidence interval).

### Outcomes averted by shielding programme

Using our counterfactual simulation where the shielding group had the same social contacts as the non-shielding group, we estimated that the shielding policy averted approximately 9100 (95% CrI: 7800-10,600) hospitalisations and 2800 (2300-3500) deaths in the shielding population (Table 1). On a relative scale, this represents a reduction of 25% (24-28%) in the number of COVID-19 hospitalisations and 23% (21-25%) in deaths compared to a counterfactual scenario with no shielding. In the whole population, we estimated a reduction of 17,700 (14,700-21,800) COVID-19 hospitalisations (13%, 12-16%) and 4500 (3700-5500) deaths (13%, 11-15%) in the scenario with shielding compared to the counterfactual scenario without shielding. In shielding people, averted outcomes are predominantly a direct result of them reducing their level of contact; while in the whole population they are an indirect effect of the lower total epidemic size due to shielding.

**Table 1.** – Projected COVID-19 severe outcomes averted by the shielding policy in England from 22 March to 1 December 2020. Rounding is to the nearest 100 for the median, and to nearest lower and upper 100 for the credible intervals (respectively).

| Severe outcome | Population | Count (95% CrI) | Percent (95% CrI) |
| --- | --- | --- | --- |
| Hospitalisations | Shielding | 9100 (7800, 10,600) | 25% (24-28) |
| Deaths | Shielding | 2800 (2300, 3500) | 23% (21-25) |
| Hospitalisations | Whole | 17,700 (14,700, 21,800) | 13% (12-16) |
| Deaths | Whole | 4500 (3700, 5500) | 13% (11-15) |

### Sensitivity analysis

The model was not sensitive to assumptions about the distribution of contacts (Supplementary Section 15), but results are sensitive to the number of social contacts (and therefore risk of infection) in the shielding group (Supplementary Section 16).

## Discussion

We estimated that between 22 March and 1 December 2020 between 7,800 and 10,600 hospitalisations and 2,300 to 3,500 deaths were directly averted in the approximately 2.25m clinically extremely vulnerable people in England, or around 25% of each outcome in the absence of shielding. We further estimated that 14,700 to 21,800 hospitalisations and 3,700 to 5,500 deaths were averted in the whole population in England, or around 13% of each outcome in the absence of shielding. Fitting a dynamic transmission model enabled us to account for time-varying and shielding– and age-specific risk of infection and captured the direct and indirect effects of shielding, giving a more complete picture of the impact of the policy. This compares with previous work, where clinical risks of COVID-19 in the general population were estimated by fitting statistical models to EHR-derived infection outcomes while using proxy estimates of the risk of infection^27^. The CoMix survey provided time-resolved age-structured contact data over the whole study period, and by using explicit information on social contact patterns in a transmission model, we were able to separate and estimate the risks of infection and of severe outcomes for the shielding and non-shielding groups.

Additionally, we estimated age and shielding-status stratified risks of becoming clinically ill and hospitalised, dying in hospital, and dying outside hospital. These risks were greater for shielding people and also increased sharply towards age 70+ at which point there was less difference in risk of death between shielding and non-shielding people. We estimated the HFR by direct calculation from a large and representative sample of the population in England, stratified by CEV status, where the outcomes are completely recorded, which adds strength to our findings. The other clinical risks were estimated using our transmission model, meaning that they were as consistent with data as possible. Key to understanding the magnitude of the indirect effects of shielding by CEV people, is the higher infectiousness of the clinically ill, whom we estimated to be 5 to 6 times more infectious than other infected people.

Supporting the validity of our model, the ONS Covid Infection Survey estimated Covid-19 prevalence from May-December is very close to our model-inferred prevalence. This agreement provides reassurance that the model estimates agree with external data sources. Our inferred parameter estimates are consistent with the estimates from existing studies^19^, although we did not use parameter values or priors from those studies, lending support to the findings.

While the 95% credible interval of the fitted model always includes the data, there are time periods where the median estimates diverge from the data, for which we have suggested possible explanations (Supplementary Section 11). For example, when recording of deaths outside of hospital they may be less likely to be tested, and there may have been improvement in clinical outcomes within hospital through time, which although we included 2 time periods in the model, could have exhibited more complex temporal patterns. Further work could investigate remaining factors.

We assumed that all CEV-identified patients shielded which agrees with ONS behavioural surveys during the first lockdown in which nearly 90% of CEV patients either self-isolated or did not have visitors^4^. However, there is imperfect alignment between being CEV and the question about risk perception in the CoMix survey. Nonetheless, the percentage of people identified through those questions agreed with the percentage of CEV-status people. People identified as CEV and notified by letter may have decreased their contacts more than those in the CoMix survey, which would mean we underestimate the impact of the policy because we overestimate the contacts of shielders. We assumed also that shielding and non-shielding groups had consistent age-specific contact patterns within those groups, which is a simplification as there is heterogeneity in the rate and age-specific assortativity of social contacts. However, this survey study remains the best available information on social contact behaviour during the pandemic in England. We also assumed that there were no regional differences in social contact, and we modelled transmission in the whole of England, which is a simplification.

We assumed that only people identified as CEV actively shielded, however other people who perceived themselves as at-risk may have decreased their contact patterns to the same degree. In addition, while there is evidence from ONS surveys^4^ that the majority of CEV people shielded, some may not have changed their contact patterns, either due to perceptions of risk or due other reasons such as their occupation or need for healthcare attendances. There is also some evidence of misclassification of CEV people that would affect our estimate, such as vulnerable people that were excluded and less vulnerable people that were included and whose shielding behaviour is unknown. These latter effects would tend to decrease the estimated impact of the policy.

The behaviour that CEV-identified people would have had without the shielding policy is unknown. We assumed they reduced their social contacts to the same extent as the general population, however their behaviour would have been a tradeoff of different influences. For instance, household members of shielding CEV people may have reduced their contacts to protect their co-habitants, which – while a small number of people – has an outsize effect on the chance of infection of the shielder. Additionally, some people may have shielded or lowered their contact rate even without the policy due to perceiving themselves at risk. In the first example, this would increase the effect of shielding, but the latter would have reduced the effect attributable to the policy. Collecting information on health-related behaviours during pandemics and epidemics is critical for quantifying the effect of policies similar to shielding.

We have estimated the impact of shielding on hospitalisations and deaths in the general population in England from 24 March to 1 December 2020. We did not include effects after this date due to complexities caused by the introduction of new variants, and differences in immunity and perception of risk caused by the introduction of vaccination. We also could not include other aspects of the shielding policy such as the impact of isolation on general wellbeing, economic aspects of the policy, and differentials in opportunities to shield due to socioeconomic or other differences in the population. Further, we could not evaluate indirect effects due to averted healthcare demand, nor could we quantify effects on people living in care homes.

## Conclusion

Our results indicate that between 24 March and 1 December 2020 the shielding policy had a major impact in reducing the burden of severe illness and death in shielding people by around 25%, and in the whole population of England by 11-15%. These large numbers are likely to have mitigated the pressure on the NHS during this period.

## Data sharing statement

All data management and analysis code is available (https://github.com/opensafely/shielding-evaluation). Data management was performed using OpenSAFELY software libraries and Python version 3.8, and analysis was carried out using R version 4.4.0, R package Rcpp version 1.0.5, and R package BayesianTools version 0.1.8. All codelists used to define data conditions and variables are openly available online at www.OpenCodelists.org for inspection and re-use.

## Author Contributions

Author contributions: Conceptualisation: RME. Design of study: JF, RME, EvL, NGD. Extraction of data: AH with input from RME, HC, JF. Contact data model: EvL with input from JF, RME. Transmission model coding and fitting: JF. Contact data and context: CJ, WJE. Contextualisation of data: LT, KBP, HJC, BM, NGD, RME. Support of OpenSAFELY platform: SB, BG, AM. Drafting manuscript: JF with RME. All authors contributed to interpretation and critical review and approved the final manuscript.

## Data Availability

The data are patient-identifiable electronic health record data and cannot be shared. A version of the data that has had privacy controls (rounding and low-number suppression) is provided in the github repository with the analysis code.

## Acknowledgments

JF was funded by the Medical Research Foundation (MRF-160-0019-ELP-EGGO-C0911) awarded to RME. RME was supported by the Medical Research Council (MR/X033260/1). K.B.P. is supported by the Medical Research Foundation (MRF-160-0017-ELP-POUW-C0909). EvL and RME are funded by the National Institute for Health and Care Research (NIHR) Health Protection Research Unit in Health Analytics & Modelling (NIHR207404), a partnership between UK Health Security Agency (UKHSA), London School of Hygiene & Tropical Medicine, and Imperial College of Science, Technology, & Medicine. The views expressed are those of the author(s) and not necessarily those of the NIHR, UKHSA, or the Department of Health and Social Care. The OpenSAFELY platform is principally funded by grants from: NHS England [2023-2025]; The Wellcome Trust (222097/Z/20/Z) [2020-2024]; MRC (MR/V015737/1) [2020-2021]. Additional contributions to OpenSAFELY have been funded by grants from: MRC via the National Core Study programme, Longitudinal Health and Wellbeing strand (MC_PC_20030, MC_PC_20059) [2020-2022] and the Data and Connectivity strand (MC_PC_20058) [2021-2022]; NIHR and MRC via the CONVALESCENCE programme (COV-LT-0009, MC_PC_20051) [2021-2024]; NHS England via the Primary Care Medicines Analytics Unit [2021-2024]. We are very grateful for all the support received from the TPP Technical Operations team throughout this work, and for generous assistance from the information governance and database teams at NHS England and the NHS England Transformation Directorate. We additionally thank Kevin van Zandvoort (LSHTM) for helpful discussions.

The views expressed in this publication are those of the authors and not necessarily those of the funders.

## Information governance and ethical approval

NHS England is the data controller of the NHS England OpenSAFELY COVID-19 Service; TPP is the data processor; all study authors using OpenSAFELY have the approval of NHS England^28^. This implementation of OpenSAFELY is hosted within the TPP environment which is accredited to the ISO 27001 information security standard and is NHS IG Toolkit compliant^29^.

Patient data has been pseudonymised for analysis and linkage using industry standard cryptographic hashing techniques; all pseudonymised datasets transmitted for linkage onto OpenSAFELY are encrypted; access to the NHS England OpenSAFELY COVID-19 service is via a virtual private network (VPN) connection; the researchers hold contracts with NHS England and only access the platform to initiate database queries and statistical models; all database activity is logged; only aggregate statistical outputs leave the platform environment following best practice for anonymisation of results such as statistical disclosure control for low cell counts^30^.

The service adheres to the obligations of the UK General Data Protection Regulation (UK GDPR) and the Data Protection Act 2018. The service previously operated under notices initially issued in February 2020 by the the Secretary of State under Regulation 3(4) of the Health Service (Control of Patient Information) Regulations 2002 (COPI Regulations), which required organisations to process confidential patient information for COVID-19 purposes; this set aside the requirement for patient consent^31^. As of 1 July 2023, the Secretary of State has requested that NHS England continue to operate the Service under the COVID-19 Directions 2020^32^. In some cases of data sharing, the common law duty of confidence is met using, for example, patient consent or support from the Health Research Authority Confidentiality Advisory Group^33^.

Taken together, these provide the legal bases to link patient datasets using the service. GP practices, which provide access to the primary care data, are required to share relevant health information to support the public health response to the pandemic, and have been informed of how the service operates.

