## Supplementary information for "Hospitalisation and mortality impact of shielding during 2020 in England: a transmission modelling evaluation using the OpenSAFELY platform"

### Supplementary Information: Methods

#### 1. Severe outcome data sources and study population

The demographic and regional distributions of the cohort are in Table S1.

**Table S1.** **Demographic and geographic characteristics of the cohort.** Age distribution is compared with that reported by ONS for England in mid 2020 ^1^. Values are shown to 1 decimal point, except the final column which is the percentage of the whole study population in the shielding group. Column 5 is derived by multiplying column 4 (as a proportion) by the study population age distribution. The total population is 23.97 million.

| Age band (yrs) | Cohort | England | % shielding in age group | % shielding in study  population | NHS region | Patients (%) |
| --- | --- | --- | --- | --- | --- | --- |
| 0 - 4 | 4.3% | 4.7% | 0.7% | 0.030% | East | 5,535,290 (23.1%) |
| 5 - 11 | 8.3% | 8.8% | 0.7% | 0.058% | East Midlands | 4,135,300 (17.3%) |
| 12 - 17 | 6.7% | 7.0% | 0.8% | 0.054% | Yorkshire and The Humber | 3,417,590 (14.3%) |
| 18 - 29 | 15.4% | 15.2% | 1.1% | 0.169% | South West | 3,365,900  (14%) |
| 30 - 39 | 14.3% | 13.5% | 1.6% | 0.229% | North West | 2,097,870  (8.8%) |
| 40 - 49 | 13.1% | 12.7% | 2.8% | 0.367% | London | 1,778,190  (7.4%) |
| 50 - 59 | 13.7% | 13.7% | 4.8% | 0.658% | South East | 1,513,190  (6.3%) |
| 60 - 69 | 10.7% | 10.7% | 8.2% | 0.877% | North East | 1,132,970  (4.7%) |
| 70+ | 13.5% | 13.7% | 13.7% | 1.850% | West Midlands | 995,370  (4.2%) |

Further to the main text, we provide further information about the EHRs accessed through OpenSAFELY and the extracted datasets on COVID-19-associated severe outcomes.

Codelists available on OpenSAFELY ^2^ were used to identify the association of COVID-19 with severe outcomes; specifically, ICD10 codes were used to identify COVID-19 to hospital admissions, and ICD10 codes U071 and U072 were used to identify COVID-19 as confirmed or suspected cause of death in the Office for National Statistics (ONS) death registry records. These severe outcomes (Table S2) were aggregated by week, and stratified by age group (1 to 9, Table S1) and shielding status (yes/no, main text Methods). Death events were further split into deaths in hospital and outside hospital (i.e. patients that died from COVID-19 without having been admitted to hospital). The three categories of severe outcome, led to 54 (3 x 9 x 2) time series of stratified weekly incidence that were used to fit the transmission model and to estimate similarly stratified probabilities of developing severe outcomes. Among the study population of 23.97 million, the number of patients with COVID-19-associated hospitalisation or death events, and the number of hospital first admissions and readmissions are shown in Table S2. Hospital readmission events were not included in the severe outcome time series and in the dynamics of the transmission model due to their relatively low frequency (9.4% of admissions) and differing interpretation from first admission, and also due to the added complexity of estimating further clinical parameters and defining readmission events in the model. The ability to differentiate readmission from first admission benefitted from EHRs being patient-level data.

**Table S2. Severe outcomes associated with COVID-19 from 1 January 2020 to 01 December 2020 in the study population (patients registered in NHS England general practices with TPP-managed EHRs**). Hospital readmissions (9.4%) were not included in the severe outcome data fitted and in the model states. Rounding: counts: 5; percentages: one decimal.

| **Patients with hospitalisation or death events** | Total |  | 55,020 |
| --- | --- | --- | --- |
|  | % by age group | 0 - 4 | 0.4 |
|  |  | 5 - 11 | 0.4 |
|  |  | 12 - 17 | 0.7 |
|  |  | 18 - 29 | 3.7 |
|  |  | 30 - 39 | 5.6 |
|  |  | 40 - 49 | 8.3 |
|  |  | 50 - 59 | 14.0 |
|  |  | 60 - 69 | 15.7 |
|  |  | 70+ | 51.2 |
| **Patient deaths** | Total |  | 16,940 |
|  | Outside hospital |  | 4,250 |
|  | % deaths |  | 25.1 |
| **Patients hospitalised** | Total |  | 50,770 |
|  | % by age group | 0 - 4 | 0.4 |
|  |  | 5 - 11 | 0.5 |
|  |  | 12 - 17 | 0.8 |
|  |  | 18 - 29 | 4.0 |
|  |  | 30 - 39 | 6.0 |
|  |  | 40 - 49 | 8.9 |
|  |  | 50 - 59 | 14.8 |
|  |  | 60 - 69 | 16.4 |
|  |  | 70+ | 48.4 |
|  | Died |  | 12,690 |
|  |  | % hospitalised | 25% |
|  |  | % deaths | 74.9% |
|  | Recovered |  | 38,080 |
|  |  | % hospitalised | 75% |
|  | Readmitted |  | 4,590 |
|  |  | % hospitalised | 9% |
| **Hospital admissions** | Single or first per patient |  | 50,770 |
|  | Readmissions (2^nd^, 3^rd^ to 6^th^) |  | 5,280 (4590, 690) |
|  |  | % admissions | 9.4% |

Statistical disclosure control in OpenSAFELY requires that no patient counts lower than 8 are outputted from the secure platform or shown in any table or figure. Cohort and aggregated counts are rounded to the nearest 5, percentage statistics are rounded to one or two digits, and ratios of counts have midpoint 6 rounding (rounded to the nearest higher multiple of 6, then decreased by 3 [17]) applied to both counts

#### 2. Directly derived clinical parameters

We assumed that the severe outcome time series extracted from the EHRs contained all the COVID-19-associated severe outcomes that occurred in the cohort during the study period, i.e. that under-reporting in the EHRs was negligible. Hence, we calculated directly the proportion of hospitalisations resulting in death rather than recovery, i.e. the hospitalisation fatality risk (HFR) stratified by age and shielding groups (Table 2), which we input into the transmission model. We also calculated a version of HFR stratified by age group only (Table S3). We further calculated the average time from hospital admission to recovery or death (Table S4), which we input into the transmission model. This calculation benefited from the EHRs being individual-based data, where individual dates of hospital admission and dates of discharge or death could be used to calculate the respective durations of hospital stay. The assumption of no underreporting was also used in the likelihood function for fitting the model to the data (Sec. 8, ‘Bayesian fitting’). The drop in HFR in the second half of the study period is likely due to improvements in treatment of clinical symptoms ^3^.

**Table S3. Hospitalisation fatality risk (%) stratified by age from 27 January to 01 December 2020.** Midpoint 6 rounding was applied to numerator (number of deaths) and denominator (number of hospital admissions). Age distribution is given in Table S1.

| Age band (yrs) | whole period | Until 30 June 2020 | From 1 July 2020 |
| --- | --- | --- | --- |
| 0 - 4 | 0.0 | 0.0 | 0.0 |
| 5 - 11 | 1.3 | 0.0 | 2.4 |
| 12 - 17 | 0.8 | 2.0 | 0.0 |
| 18 - 29 | 0.4 | 1.1 | 0.2 |
| 30 - 39 | 1.9 | 2.3 | 1.3 |
| 40 - 49 | 4.9 | 6.0 | 3.5 |
| 50 - 59 | 9.4 | 12.8 | 5.6 |
| 60 - 69 | 19.2 | 25.1 | 12.8 |
| 70+ | 41.1 | 48.4 | 31.7 |

**Table S4. Average time to outcomes associated with COVID-19 while in hospital.** Time to recovery: since admission to discharge, or, for patients with multiple admissions, since the first admission to first discharge. Time to death: since admission or, for patients with multiple admissions, since the first admission. Durations are averaged over the cohort, with no differentiation between the duration of clinical development in shielding and non-shielding patients; such a differentiation is already made through the hospitalisation fatality risk (Table S3) and both quantities are multiplied within the transmission model. Continuous variables do not require specific rounding for output release.

| Mean (median) (days) | whole period | Until 30 June 2020 | From 1 July2020 |
| --- | --- | --- | --- |
| Time to recovery | 12.0 (6) | 10.1 (6) | 14.2 (7) |
| Time to death | 13.9 (10) | 12.5 (9) | 16.8 (12) |

#### 3. Transmission model

The deterministic implementation of the compartmental transmission model (Figures 1 and S1), was made through the following ordinary differential equations (see main text ‘Transmission model’ and Tables S5-S6 for state variable and parameter notation; note that each rate parameter is the inverse of the corresponding average period):

$\frac{dS_{i,s}}{dt}= - l_{i,s}S_{i,s}$	(1)

$\frac{dE_{i,s}}{dt}= l_{i,s}S_{i,s}-[{r_{EI} y}_{i,s} {+ r_{EU}{(1-y}_{i,s}) ] E}_{i,s}$	(2)

$\frac{dU_{i,s}}{dt}= {r_{EU}{(1-y}_{i,s}) E}_{i,s}-{r_{UR} U}_{i,s}$	(3)

$\frac{dI_{i,s}}{dt}= {r_{EI}y_{i,s} E}_{i,s}-{{[r_{IR} \left( 1-h_{i,s}-d_{i,s} \right)+ r_{IH} h_{i,s}+ r}_{IO} d_{i,s} ] I}_{i,s}$	(4)

$\frac{dR_{i,s}}{dt}= {r_{UR} U}_{i,s}+{r_{IR} \left( 1-h_{i,s}-d_{i,s} \right) I}_{i,s}+ {r_{HR} \left( 1-m_{i,s} \right) H}_{i,s}$	(5)

$\frac{dH_{1,i,s}}{dt}= {r_{IH} h_{i,s} I}_{i,s}-r_{H}H_{1,i,s},\ldots,$

$\frac{dH_{n,i,s}}{dt}={r_{H} H}_{n-1,i,s}-{n [ r}_{HR}{\left( t \right) (1-m_{i,s}\left( t \right))+r_{HD}(t) m_{i,s}(t) ] H}_{n,i,s}$	(6)

$\frac{dO_{1,i,s}}{dt}= {r_{IO} d_{i,s} I}_{i,s}-{n r}_{OD}O_{1,i,s},\ldots, \frac{dO_{n,i,s}}{dt}={{n r}_{OD} O}_{n-1,i,s}-{n r}_{OD}O_{n,i,s}$	(7)

$\frac{dD_{H,i,s}}{dt}= n r_{HD}{\left( t \right)m_{i,s}\left( t \right) H}_{n,i,s}$	(8)

$\frac{dD_{O,i,s}}{dt}= n r_{OD}O_{n,i,s}$,	(9)

where, *i* = 1…9 and *s* = 0 or 1 indicate age and shielding-status strata (no or yes); *r_H_ = n r_HD_(t)*; *n =* 5 and *r_HD_(t)* and *r_HR_(t)* change value on 30 June 2020; *t* is continuous time during the study period. *D_H,i,s_(t)* and *D_O,i,s_* are the cumulative numbers of deaths in hospital and outside hospital. Data were fitted to the model weekly incidences of hospitalisation and deaths obtained by integrating the rate of change of the variables over each week.

After an average latent period, exposed individuals with age *i* and shielding status *s* became clinically infected (*I*, displaying clinical symptoms) or sub-clinically infected (*U,* with few or no symptoms, and thus unascertained) with respective probabilities *y_is_* and *1-y_is_*. We assumed there was no waning of immunity to re-infection during the study period. Clinically infected individuals developed severe COVID-19 symptoms and either became hospitalised (*H*) after an average hospital-admission period, or died in the community (*D*) after an average outside-hospital mortality period, or recovered (R) after the average infectious period, with respective probabilities *h_is_*, *d_is_* and *1-h_is_-d_is_*. Hospitalised individuals, either progressed to recovery after an average hospital-recovery period, or died after an average hospital-mortality period, with respective probabilities 1*-m_is_* and *m_is_*. For ease, we called the probabilities *h*, *y*, *m*, *d* ‘clinical responses’ as they are risks of development of clinical stages during SARS-CoV-2 infection.

The force of infection (rate at which a susceptible individual becomes exposed) is given by:

$l_{i,s}=\beta u_{i} \sum_{j} c_{ij,s, t}\left[ p_{sh,j} \frac{I_{j,1}+f U_{j,1}}{N_{j,1}} + \left( 1-p_{sh,j} \right) \frac{I_{j,0}+f U_{j,0}}{N_{j,0}} \right]$	(10)

where *c_i j,s,t_* (Table S5) is the daily contact rate of an individual of age *i* and shielding status *s* with individuals of age *j* (of any shielding status) at time *t*; and p*_sh,i_* is the age-specific probability of shielding (Table S1). For simplicity, vital dynamics (natural deaths, births, and ageing) are not included in each compartment; we assumed ^4–6^ that the effects of these processes could be neglected during the short duration of the study period.

The equations were solved numerically via Euler time steps; to ensure accuracy, each step was 0.1 day, which is no larger than 1/10^th^ of the shortest time scales associated with the model rate parameters, both their prior lower bounds (Figure S1) and their estimates (Table S5). Testing indicated this approximation had 98% to 93% accuracy, while computation was tens of fold faster compared to alternative numeric implementations for MCMC sampling.

#### 4. Model parameters

All parameters of the transmission model are defined in Table S5. Here we add further to the description of model parameters in the main text.

Some of the initial clinical infections that seeded the modelled epidemic led to the initial hospitalisations. Hence, we set the age distribution of the estimated total initial infections, *N_I0_*, based on the age distribution of hospitalisation incidence observed at the start of the modelled epidemic period.

Substantial reduction in dimension of the parameter sampling space was achieved by using functional forms for the clinical response probabilities *y_i,s_*, *h_i,s_*, *d_i,s_*, rather than estimating each response for each of the 9 age groups. We assumed that each response has an exponential relationship to age characterised by shielding-status specific amplitude and rate parameters. For example, the probability of hospitalisation is *h_i_(a_i_) = h_A,s_ exp(h_r,s_ a_i_)*, where a_i_ is the mid-point age of group *i*, and *h_A,s_* and *h_r,s_* are the amplitude and rate parameters for shielding status *s*. Such a two-parameter relationship is supported by various studies ^7,8^. In the case of the clinical fraction *y*, previous estimates for the general population (not shielding stratified) were higher in the two youngest than the in the mid-age groups ^9^, hence we took the two younger-age *y* values to be as previously estimated and applied the exponential relationship to the remaining age groups. This functional approach required estimation of 12 parameters (3 outcomes x 2 curve parameters x 2 shielding strata) to estimate the clinical responses *y*, *h*, *d*, as opposed to 50 parameters otherwise.

In the case of deaths in hospital, the probabilities *m* could be calculated directly as proportions among the EHRs, as we expect the EHRs to have negligible missing data or under-reporting (Sec. 2, ‘Directly derived clinical parameters’). The average durations from hospital admission to death and from hospital admission to recovery could also be directly estimated from the EHRs, from which individual-level length of hospital stay could be obtained. We estimated these durations (Table S4) and *m* (Table 2) for each of two periods, before and after 30 June 2020 (the midpoint of the study period), to capture change in severe outcome development in hospital as clinical treatment improved during the pandemic ^3^.

Note that the probabilities *y*, *h*, m*,* *d* relate to widely used epidemiological indicators. As already stated, *m* is the HFR (by shielding status in Table 1 and for the whole population in Table S3), and the product *y h* is the infection hospitalisation risk (IHR). Furthermore, the infection fatality risk (IFR) is given by *y ( h m + d)*.

The force of infection *λ_i,s_* on a susceptible *i*, *s* in week *w* (Sec 3, ‘Transmission model’) includes social contact with the infectious individuals in every age and shielding-status group in week *w*. The social contact rates are based on data from the CoMix survey conducted during the first year of the pandemic ^10^, and are temporally-varying due to behavioural changes such as compliance with lockdowns and other influences.

For the exposed individuals, an additional pathway was added to the model (further to the pathway from exposed to infected compartments) to predict the number of individuals that would test positive in a PCR test. This separation was necessary because positivity is expected to last longer than the infected state; we used a previously estimated duration of ^11,12^ (Table S5). We used Erlang distributions with five compartments to represent (with reduced left-skewness) the duration of positivity, the duration from admission to death in hospital, and the duration from clinical infection to death outside hospital.

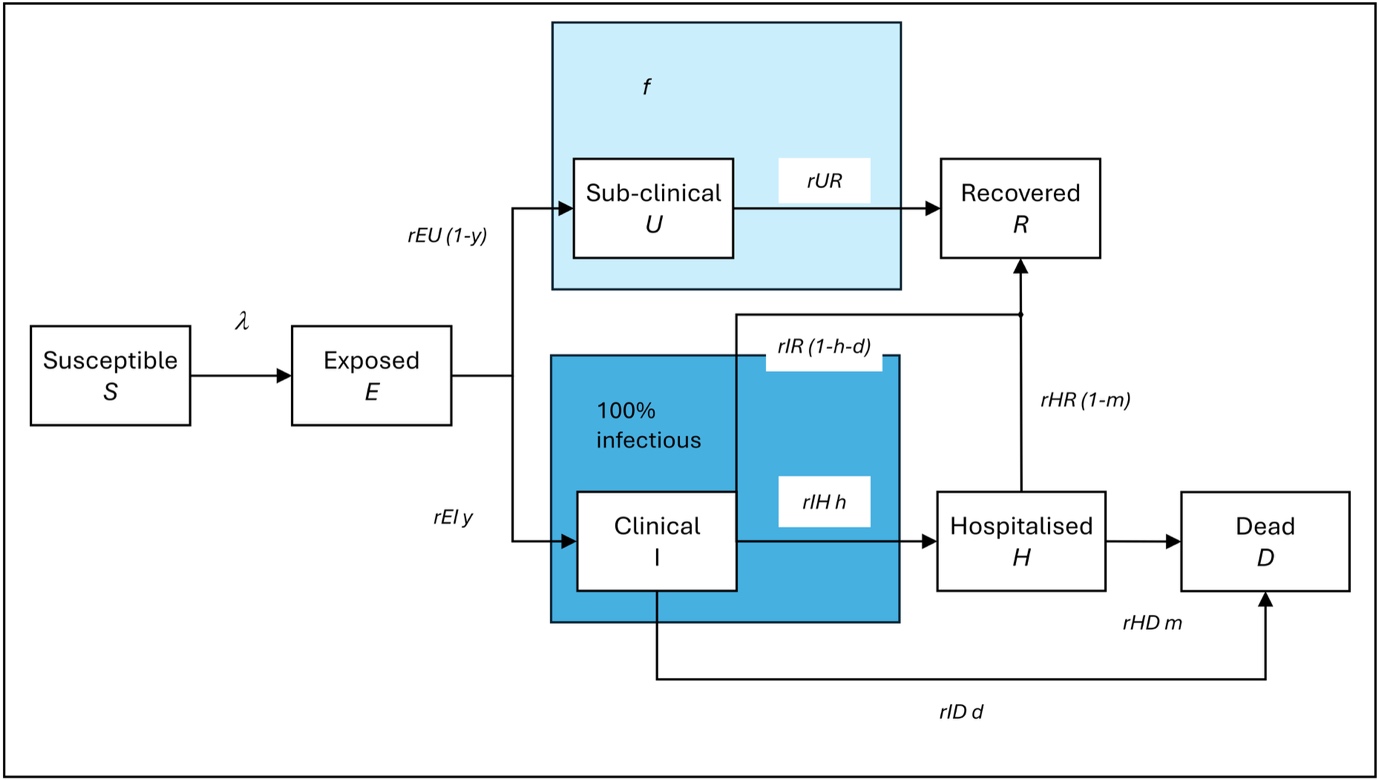
**Figure S1. Model diagram with symbols.**

**Table S5. Parameters of the transmission model.** Parameters of the model that depend on age and shielding status have indices *i* and *s*. Notation: an average duration or period is the inverse of the corresponding transition rate, e.g. *t_EI_ =1/r_EI_*, where *r_EI_* is used directly in the model. Parameter *t_ID_* was not directly sampled via MCMC; each *t_ID_* posterior value is the sum of jointly sampled *t_IH_* and *t_OD_* values. HFR hospitalisation fatality risk. Parameter *p_I0_* was estimated for a given fixed value of parameter *p_E0_* as the two parameters were highly correlated; we chose a fixed value with the lowest DIC among a given set.

| **Symbol** | **Description** | **Specific (age, shielding)** | **Value** | **Source/method** |
| --- | --- | --- | --- | --- |
| *t_EI_, t_EU_* | Latent period | none | Estimated | *t_EU =_ t_EI,,_* |
| *t_IR_, t_UR_* | Infectious period of clinical infections | none | Estimated |  |
| *t_UR_* | Infectious period of sub-clinical infections | none | Estimated |  |
| *R_0_* | Basic reproduction number | NA | Estimated |  |
| *p_I0_* | Proportion of infected people on 27 January 2020 | none | Estimated |  |
| *p_E0_* | Proportion of exposed people on 27 January 2020 | none | 1000/ 56,430,000 | Assumption  with lowest DIC among 500, 750, 1000, 1500 |
| *t_IH_, t_IO_* | Period to hospital admission from onset of clinical infection | none | Equal to the estimated *t_IR_* | ^5^ |
| *t_HR_* | Period to recovery in hospital, from admission to discharge | age | Calculated | EHR data |
| *t_HD_* | Period to mortality in hospital, from admission to death | age | Calculated | EHR data |
| *t_ID_* | Period to mortality outside hospital from onset of clinical infection | none | *t_IO_ + t_OD_*  averaged over samples | Calculated |
| *t_OD_* | Period to mortality outside hospital from needing hospital admission | none | Estimated |  |
| *t_C_* | Duration of positivity in PCR test upon infection | none | 8.5 d | ^11,12^ |
| *f* | Infectiousness of unascertained infection in relation to clinical infections | none | Estimated |  |
| *β* | Probability of infection on contact | none | Estimated |  |
| *y_i,s_* | Probability of clinical symptoms in an infection (clinical fraction) | Age, shielding | Estimated | Bayesian inference;  exponential function of age for age groups 3 to 9; values for age groups 1-2 are from literature [16] |
| *h_i,s_* | Probability of hospital admission while clinically infected | Age, shielding | Estimated | Bayesian inference,  exponential function of age |
| *m_i,s_* | Probability of death while in hospital, HFR | Age, shielding | Calculated | EHR data |
| *d_i,s_* | Probability of death outside hospital while clinically infected | Age, shielding | Estimated | Exponential function of age |
| *u_i_* | Susceptibility to infection | Age | 0.40, 0.39, 0.38, 0.72, 0.86, 0.80, 0.82, 0.88, 0.74 | For the age groups in Table 1. Derived from ^9^ by adjusting to the current age groups. |
| *k_H_* | Overdispersion of hospitalisation events | none | Estimated | Negative binomial distribution at each time point |
| *k_D_* | Overdispersion of death events | none | Estimated | Negative binomial distribution at each time point |
| *c_i,j,s,t_* | Contacts per day of a person age *i* and shielding status *s* with persons age *j* at time *t* | Age, shielding | Derived, see ‘Social contact survey data’ | CoMix social contact survey ^10^ and regression modelling |

It was not possible to jointly estimate *p_I0_* and *p_E0_* due to non-identifiability, so we tested 4 different values for *p_E0_* while estimating *p_I0_*, and used 1000 as it gave the lowest DIC of tested values.

#### 5. Social contact survey data

We input into the transmission model age-structured and shielding-status-specific contact matrices generated by a regression model fitted to social contact data from the CoMix survey [12]. We implemented this regression to extrapolate the data to the non-surveyed period of 27 January – 23 March 2020; to address a two month delay in the collection of contact data for children; and to smooth out some extreme outliers in the data. We extracted from the survey their biweekly estimates of the mean daily contacts of residents in England stratified by the age groups of the participant and contact persons. For simplicity, we chose the age groups in the transmission model and EHR data analysis (Table 1) to be the same as these in the CoMix survey. As in the published CoMix analysis ^10^, we censored contacts to 50 per participant per day to avoid disproportionate contribution from a few outliers. As with the severe outcome data, we aggregated the social contact data at national level.

To identify which survey participants where potentially shielding, we used information about the participant’s perception of risk as a proxy. The specific questions (answers) were: “Are you or any other household member in a high-risk group under which the annual influenza vaccine would usually be offered by the NHS?” (Yes/No), “Are you or any other household member in a high-risk group, meaning you could have serious symptoms if you contracted Coronavirus (COVID-19)?” (Yes/No), “To what extent do you agree or disagree with each of the following statements? Coronavirus would be a serious illness for me” (Strongly agree/Tend to agree/Tend to disagree/Strongly disagree). We selected the participants that answered, respectively, “Yes”, “Yes”, and “Strongly agree”. This criterion led to an estimate of 7.9% of “shielding” participants among the total participants. However, this estimate is only indicative because, as participants responded to multiple survey rounds (4.5 on average), we assumed that their responses were consistent across rounds, while the reality is likely to have been mixed in this respect. In addition, while our proxy criterion for identifying shielding participants is likely to have picked up most if not all of those that had been identified as CEV, it is also very likely to have picked up other participants that were shielding for other reasons. Therefore, we expected to obtain a percentage of “shielding” survey participants (7.9%) greater than the estimated 4.2% shielding patients in the EHRs.

The important outcome of our proxy approach is that it allowed us to make an objective quantitative distinction between the levels of contact of shielding and non-shielding people. It was less important whether the proportion of shielding participants matched the proportion of CEV patients in England because the survey may or may not have been representative of the population of CEV-identified people in England, and because the level of contact of people shielding for other reasons may have been broadly similar. In addition, the social contact estimates obtained via the regression model (see below) showed little sensitivity to the shielding percentage and its age distribution, i.e. to whether these were inferred jointly in the regression model or were inputted as the values measured in the study population (see ‘Shielding population’); the different regression outcomes showed only minor detectable differences in the contact rates of shielding people. The same lack of sensitivity to these versions of the estimated social contacts was found in the outcomes of fitting the transmission model.

#### 6. Inferring contact data

Given that the CoMix survey started on 24 March 2020, and that children’s contact data were collected only from May 2020, we used Bayesian regression to extrapolate the contact data of each age and shielding-status group to the start date of the modelled epidemic (27 January 2020). The regression covariates were the frequencies of attendance in schools taken from school attendance data sets ^13^; visits to workplaces and to retail and recreation venues, as measured by Google mobility ^14^; and temperature ^15^. The CoMix data sets used for this analysis are publicly available ^16^. The outputs of this approach were age-structured contact matrices, for the non-shielding population and for the shielding population, for each fortnight within the 45 weeks of the modelled period (27 January to 01 December 2020). For the midpoint of each fortnight, we repeated the data of the previous week. For each population and fortnight, we took the mean over 250 sampled matrices drawn from the posterior distribution of the model given the data. Credible intervals for total age specific and overall contact rates were also estimated based on these matrices.

The shielding policy started officially on 22 March 2020 ^17,18^. We assumed that CEV-identified people were aware of their risk and gradually started to shield three weeks earlier. Hence, we assumed that the social contact matrix of shielding people had the following biweekly transition steps (where the date corresponds to the midpoint of each fortnight): on 02 March 2020, the same as for the non-shielding population; on 16 March 2020, mean of the non-shielding population and shielding population matrices projected through regression; on 30 March 2020, the shielding population matrix projected through regression. Figure 1 shows the resulting mean daily rates of social contact in the non-shielding and shielding populations, aggregated over the age groups of the participants and of their contacts.

We fitted a contact model to the CoMix data. The contact model assumed that the change of the number of contacts is linearly correlated with the explanatory variables (school attendance (from ^13^), temperature ^15^ and google mobility data on work and recreational (visit) activity (Figure S2) ^13,19^.

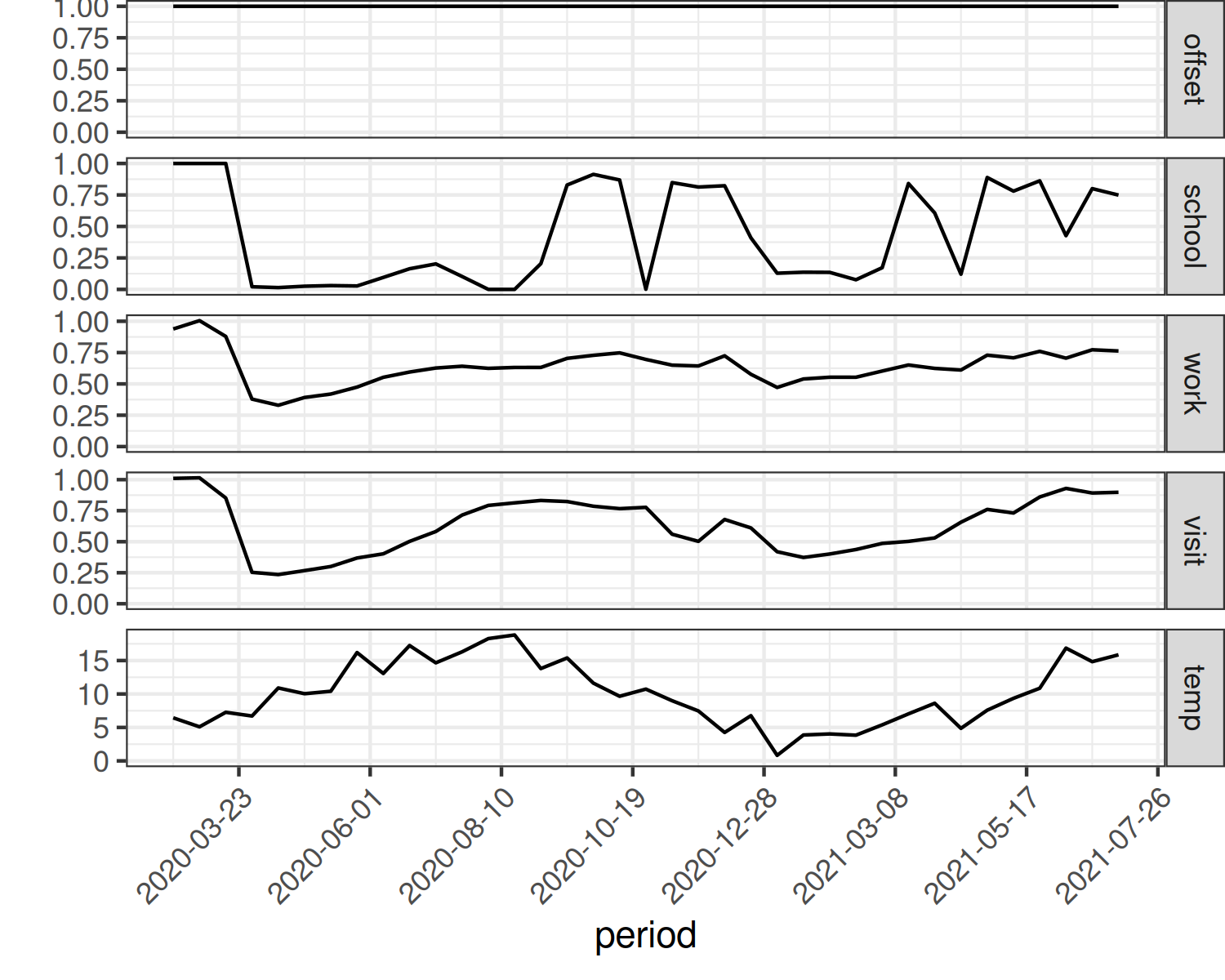

**Figure S2** **1: Time series of the explanatory variables in the contacts model.**

The contact matrix is then defined as

$$c_{ij}=\frac{a_{i}+a_{j}}{2}\hat{c}_{ij} .$$

We assume $a_{i}$ depends on Google mobility and school attendance data sets linearly, e.g.: $a_{i}=\beta_{0}+\beta_{1}x_{1}+\ldots$, where $x_{i}$ is the explanatory variable and $\beta_{i}$ is the coefficient.

The contact matrix can then be split into contact rates of the shielding and non-shielding population as follows:

$$\begin{matrix} c_{ij}^{oo} & =c_{ij} \\ c_{ij}^{os} & =c_{ij}\alpha_{j} \\ c_{ij}^{so} & =\alpha_{i}c_{ij} \\ c_{ij}^{ss} & =\alpha_{i}c_{ij}\alpha_{j} , \end{matrix}$$

where $s$ indicates a shielding individual, $o$ a non shielder (other), $c_{ij}^{xy}$ the probability of a random individual with shielding status $x$ and age group $i$ having a contact with a random individual with shielding status $y$ and age group $j$, $\alpha_{i}$ is the reduction in contact rate of a shielding individual in age group $i$ and $c_{ij}$ is the probability in the absence of shielding. These matrices are used in the theoretical derivation of the shielding and non-shielding matrices, but are not themselves recorded in data or fitted to data. They can then in principle be used to calculate the probability of a non-shielder or shielder in age group $i$ having a contact with anyone in age group $j$ ($c_{ij}^{o}$ and $c_{ij}^{s}$ respectively):

$$\begin{matrix} c_{ij}^{o} & =c_{ij}^{oo}\left( 1-p_{j} \right)+c_{ij}^{os}p_{j} \\ & =c_{ij}\left( 1-p_{j} \right)+c_{ij}\alpha_{j}p_{j} \\ & =c_{ij}\left( 1+\left( \alpha_{j}-1 \right)p_{j} \right) \\ c_{ij}^{s} & =c_{ij}^{so}\left( 1-p_{j} \right)+c_{ij}^{ss}p_{j} \\ & =\alpha_{i}c_{ij}\left( 1-p_{j} \right)+\alpha_{i}c_{ij}\alpha_{j}p_{j} \\ & =\alpha_{i}c_{ij}\left( 1+\left( \alpha_{j}-1 \right)p_{j} \right)=\alpha_{i}c_{ij}^{o} \end{matrix}$$

with $p_{j}$ the probability that someone shields in age group $j$. This probability was derived from OpenSAFELY cohort data. All other parameters are fitted.

From the fitting we can derive the number of contacts for each cohort ($c_{ij}^{s}N_{j}$ and $c_{ij}^{o}N_{j}$). Contact survey data has the number of contacts different participants have with different age groups. We can then define the following likelihood for the participants as follows:

$$\begin{matrix} \mathcal{L=}\prod_{\nu} \prod_{k} \prod_{j} P\left( y_{j}^{\nu,k};c_{k\mapsto i,j}^{\nu}N_{j} \right) \end{matrix}$$

where $\nu\in s,o$ is the shielding state of the participant, $k$ is the participant, $i$ is the age group of participant $k$ and $j$ is the age group of the contact. We assume that the number of contacts is Poisson distributed.

The fits are shown for non-shielding (Figure S3) and shielding (Figure S4) as well as coefficients of the estimation (Figure S5). Alpha parameters for the shielding population are shown in Figure S6.

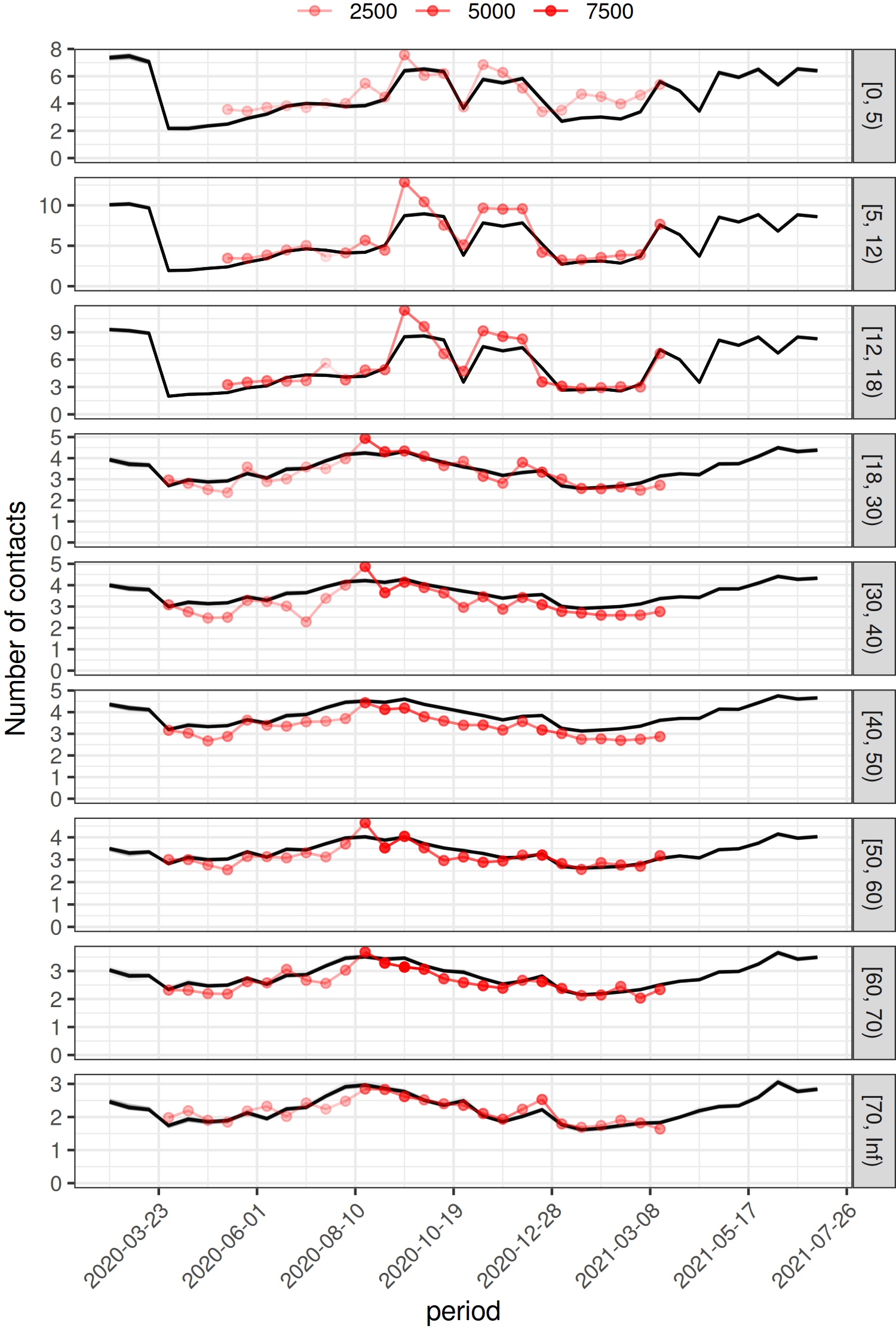

**Figure S3.** **2**Mean number of contacts over time by age group for the non-shielding cohort (black). Red is the data, with the opacity representing the total number of contacts underlying that data point.

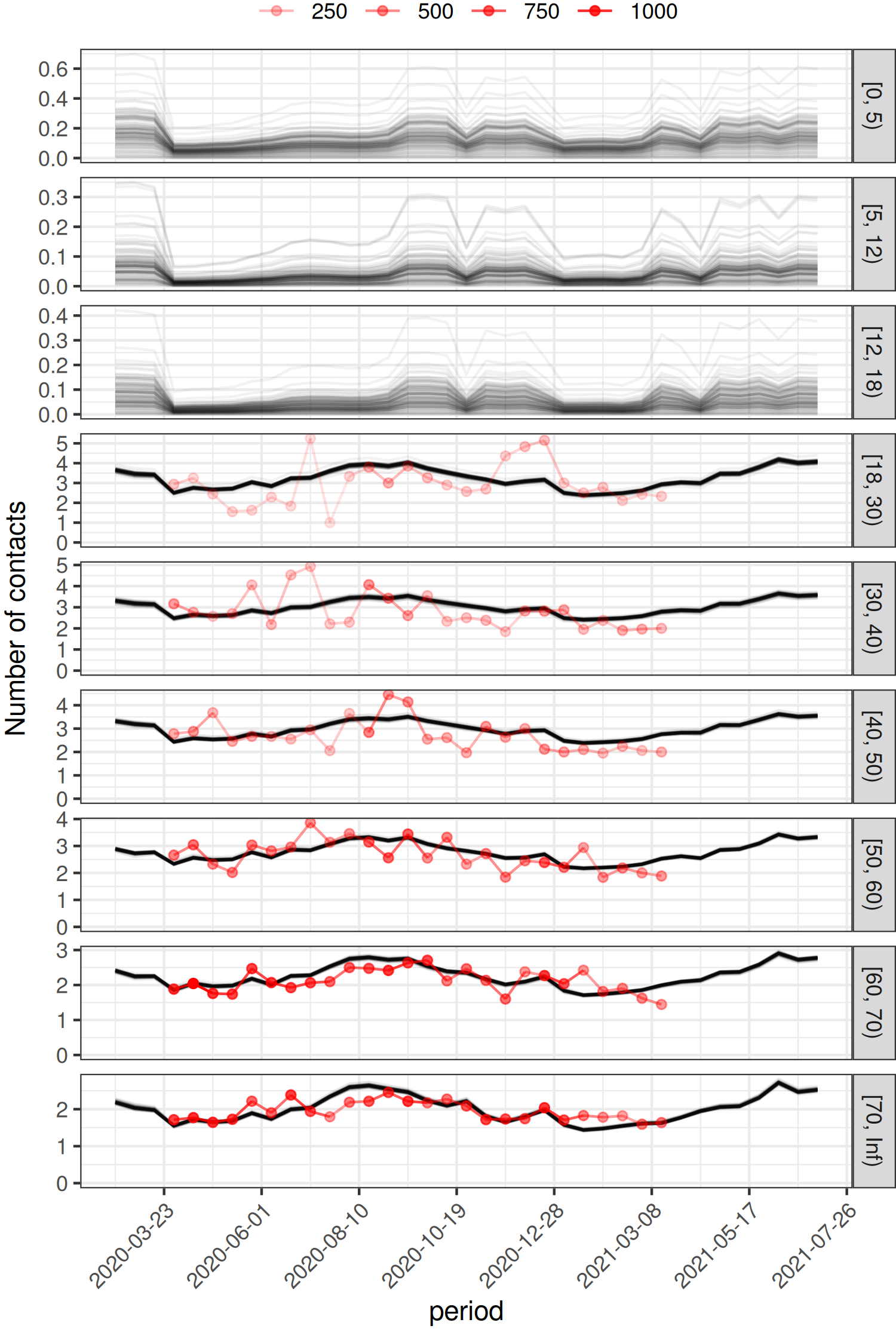

**Figure** **S43:** Mean number of contacts over time by age group for the shielding cohort (black). Red is the data, with the opacity representing the total number of contacts underlying that data point.

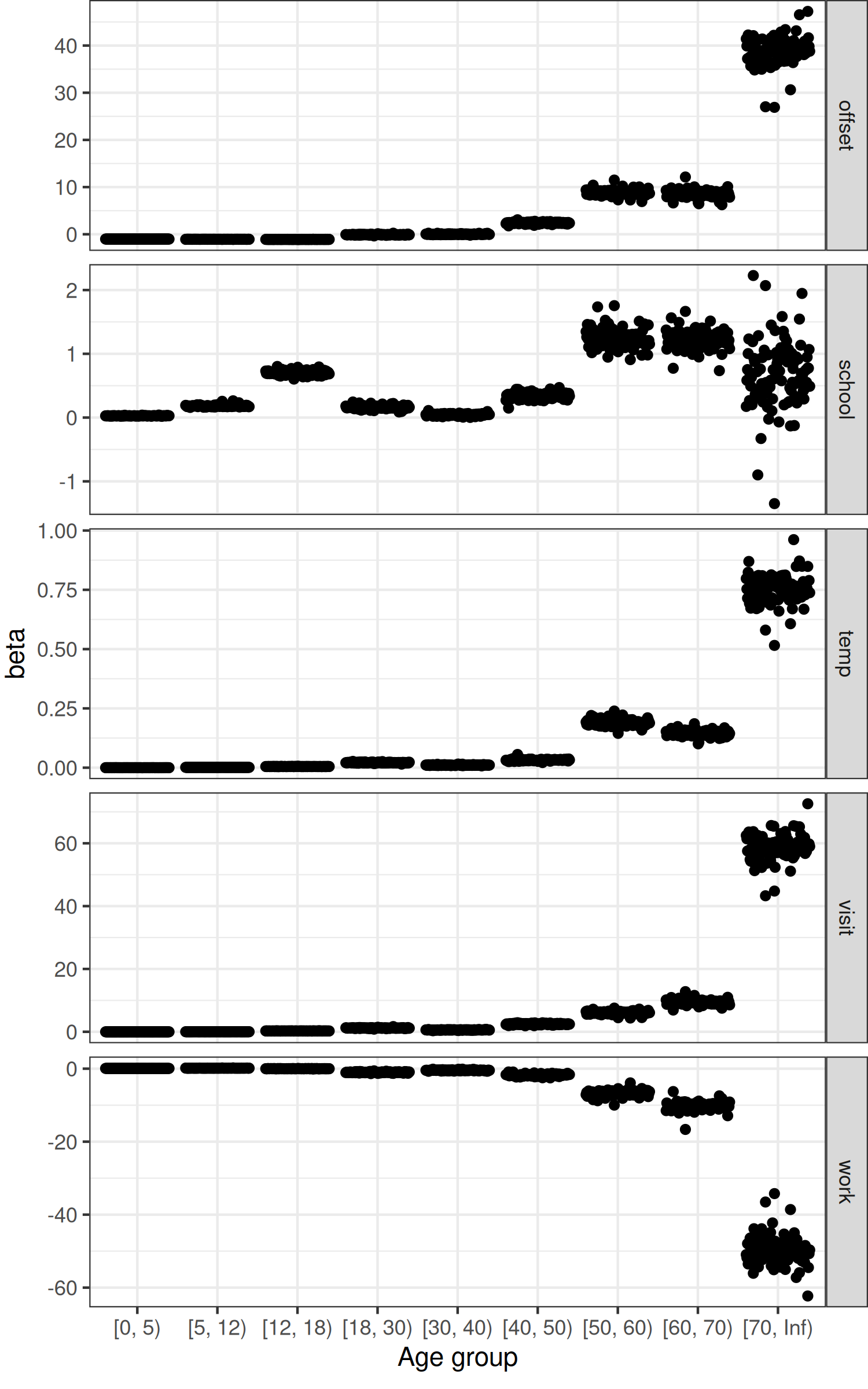

**Figure S5** **4:** Coefficients for the explanatory values by age group.

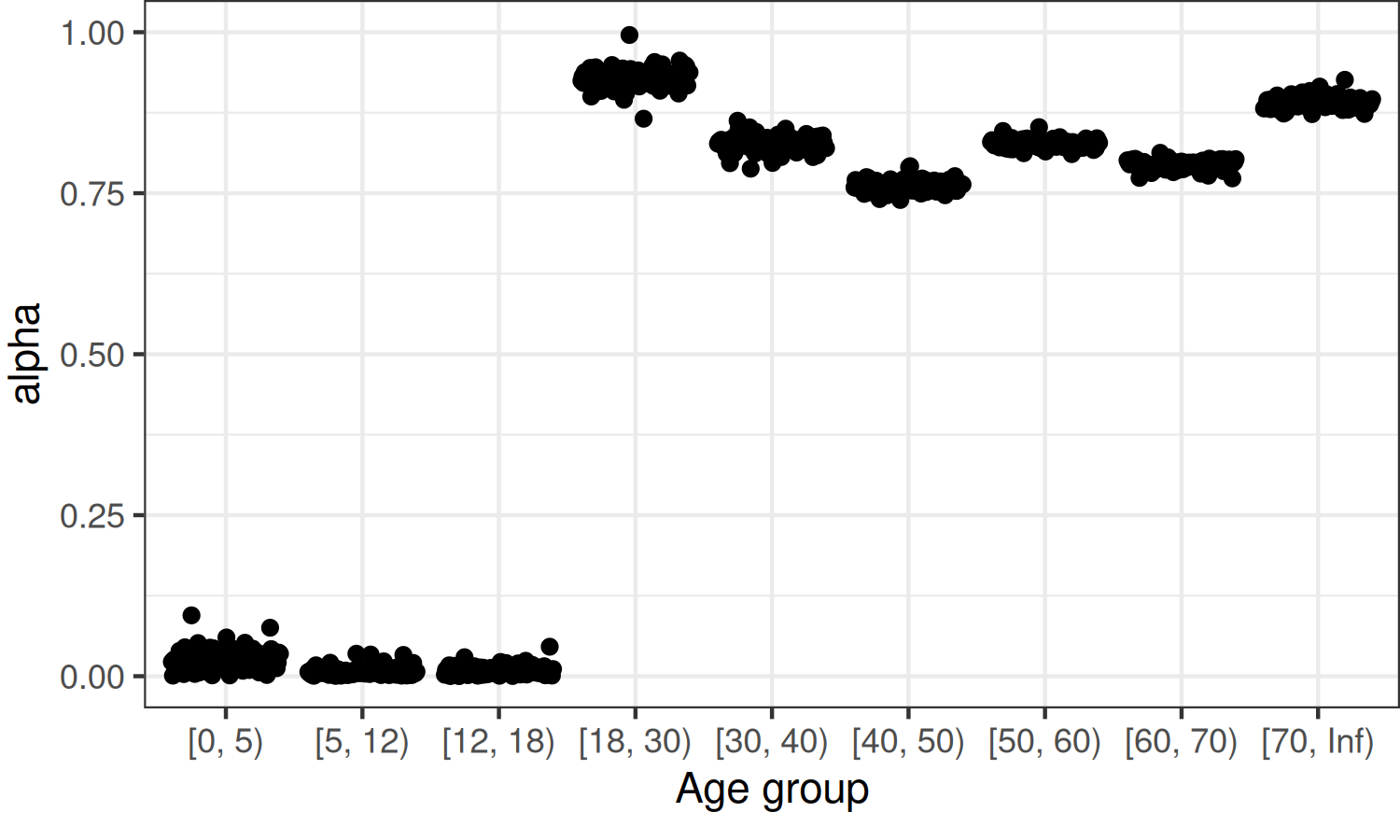

**Figure S6** **5:** Relative contact rates in the shielding population (alpha).

#### 7. Basic reproduction number

The basic reproductive number (*R_0_*), defined as the average number of secondary infections caused by a single infectious individual in an otherwise susceptible population, is calculated using the next generation matrix (NGM) of the transmission model ^20^, whose *i*^th^ row and *j*^th^ column, at time *t*, is (see Sec. 3, ‘Transmission model’ and Table S5 for notation):

$NGM_{ij,t}=\beta u_{j} c_{i,j,0,t} \left[ y_{i,0} d_{I,i}+f\left( 1-y_{i,0} \right) d_{U} \right] ,$	(11)

$d_{U}=\frac{1}{r_{UR}},$

$d_{I,i}=\frac{1}{r_{IR}\left( 1-h_{i,0}-d_{i,0} \right) + r_{IH}h_{i,0}+r_{IO}d_{i,0}}$ ,	(12)

where *c_ij,0,t_* (Table S5) is the daily contact rate of an individual of age *j* and not shielding (*s*=0) with individuals of age *i* (of any shielding status) at time *t*; *β* is the probability of effective transmission by an infectious contact; *u_i_* is the susceptibility at age *j*; d*_U_* is the average duration of sub-clinical infectiousness; and d*_I,i_* is the average duration of clinical infectiousness at age *i* (i.e. while not becoming hospitalised, at rate *r_IH_ h_j_*, or in need of hospitalisation but not being admitted to hospital, at rate *r_IO_ d_i_*). *R_0_* is the absolute value of the dominant eigenvalue of the NGM.

We defined *R_0_* via the NGM for the larger, non-shielding population, which comprised 96% of the population in England, and therefore made the principal contribution to contacts in *R_0_*. We assumed that the contribution of the shielding population would have a negligible impact on the *R_0_* calculation. Using the NGM for the whole population would have doubled its dimension and considerably increase the computation time needed to calculate the NGM eigenvalues. The pre-pandemic *R_0_* was obtained using the earliest weeks of social contact data when contacts rates did not change over time (27 January to 22 February 2020, Figure 1). As the social contacts (based on the CoMix survey ^10^) varied over the study period up to 01 December 2020, so did *R_0_* (Figure S11). No account is taken during this period for change in population immunity ^4^ as it was deemed low ^5^, hence we do not use the term effective reproductive number for the time-varying *R_0_*.

#### 8. Bayesian fitting

We parameterised the transmission model and made inferences using Bayesian methods. Specifically, we sampled the model joint posterior distribution (given the data) using the Differential Evolution (DE) ^21^ implementation of the Metropolis-Hastings MCMC algorithm ^22^. The implementation was further optimised through the use of of snooker parameter proposals and past-event updates ^23^, which allow DE to perform with as few as 3 independent sampling chains in highly-dimensional sampling spaces. We implemented this approach in the R package BayesianTools ^24^, version 0.1.8, using its DEzs algorithm option and the defaults of 3 sampling chains and of a multi-dimensional normal density centred on the current iteration values to propose updates to the joint posterior parameter distribution. MCMC sampling was carried out over 400,000 iterations equally distributed among the chains, of which half were discarded as burn-in, and to which 1/7 thinning was applied, resulting in 3 chain samples over 9,500 long.

The sampling space comprised 21 model parameters (Table S5). Beta prior distributions (with both shape parameters equal to 4) were used for each parameter with bounds defined by biological plausibility (Figure S7); the Beta prior option is a built-in option in the above package. One parameter (*p_I0_*, Table S6) was log-transformed, and the two dispersion parameters used in the likelihood were pareto transformed to manage the high sensitivity to the untransformed parameters; the other parameters were not transformed.

We used Bayesian Evidence Synthesis ^25^, jointly fitting the weekly incidence of the three COVID-19-associated severe outcomes (hospitalisation, death in hospital, and death outside hospital) over 45 weeks (27 January to 01 December 2020). Where possible, for each severe outcome, we fitted the count time series of each of the 9 age groups and each of the two shielding-status groups. To tackle fitting of the model to scarce events, for each severe outcome and age and shielding-status group whose counts were too low (i.e. exhibited no clear temporal pattern), we fitted the event’s total count over the study period rather than the counts at every time point. We fitted time series for age groups 5-9 (Hospitalisations), and 7-9 (Deaths in hospital, and Deaths outside hospital). For other groups we fitted total counts.

Each time series aggregated counts from TPP-served GPs across all 9 NHS England regions; this decision to aggregate regions was based on the national scarcity of weekly events in many strata, and their even greater scarcity regionally. As the model predicted outcomes in England, each time series was scaled-up by the TPP-to-national demography ratios (around 2.5) for each age group. Here, we assumed that the epidemic across the remaining English population was similar to that in the cohort of patients registered in TPP-served GPs, which distribute across all NHS England regions (Table S1); this assumption is backed by analyses that show these practices are representative of the national population ^26^.

As likelihood of the time series data, given the model, we used a negative binomial density with mean equal to the model predicted weekly incidence of each severe outcome and size parameter equal to the overdispersion parameter for either hospitalisation or death events (*k_H_* and *k_D_*). The likelihood makes no account for under-reporting in the observation process because we assumed that the EHRs completely reported all clinical events.

To generate model outputs, the unknown parameter *β* (Table S5 and Sec. 7, ‘Basic reproduction number’) was calculated as the value of *R_0_* divided by the absolute value of the dominant eigenvalue of NGM’, where NGM’ is the next generation matrix NGM without *β* as a factor. *R_0_* was directly estimated, rather than *β*, because there was greater prior knowledge about *R_0_*. At each step of the MCMC estimation, values for the estimated parameters were proposed and then fed into the NGM’ to calculate *β* and into the model to calculate the incidence of severe outcomes, from which the data likelihood was evaluated and the parameter proposal was either accepted or rejected.

Parameter posterior distributions were characterised by their median and 95% credible interval (95% CrI). Posterior predictive distributions, obtained via simulation from the posterior samples, were used for comparison with observation data and were characterised by their median and 95% prediction interval (95% PI). MCMC convergence and mixing were assessed through multiple diagnoses: inspection of the parameter marginal posterior densities (Figure S7) and their traces (Figure S8), the Gelman-Rubin potential scale reduction factor (PSRF) ^27^, the effective sample size, and parameter Pearson pair correlations (data not shown).

#### 9. COVID-19 positivity survey data

We used publicly available virus positivity data for England from the ONS Coronavirus Infection Survey ^28^ to test the transmission model parameterised on the EHRs. The survey used nose and throat swabs from survey participants. The swabs were processed in the laboratory for the presence of SARS-CoV-2 using real-time reverse transcriptase polymerase chain reaction (RT-PCR) testing. Based on the test results, ONS estimated the average number of new positive test cases per week, and how many people across England (and the other UK nations) would have tested positive for a COVID-19 infection, regardless of whether they report experiencing symptoms ^29^. The data are displayed in Figure 4.

We downloaded file “covid19infectionsurveydatasetsengland20230310.xlsx” ^28^, copied sheet “UK summary – positivity”, and extracted columns “England Time period”, “England Estimated average % of the population testing positive for COVID-19”, “England 95% Lower confidence/credible interval”, “England 95% Upper confidence/credible interval”. As each weekly estimate of positivity was a rolling-average of the previous two weeks, we took the midpoint of each two-week period as the date corresponding to each positivity record. The dates ranged from 03 May 2020 and beyond the end of our study period.

### Supplementary Information: Results

#### 10. Estimated model parameters and MCMC diagnostics

The parameters characterising the hospital pathway were directly estimated from the EHRs (Table S5), i.e. the duration of the periods from hospital admission to recovery and to death (Tables S4 and S6) and the probability *m* of mortality in hospital for each half of the study period (before and after 30 June 2020) (Table S6). The probability *m* (Figure S10 i-j) appears to follow an exponential relationship with age for each shielding status and for each half of the study period, similarly to our assumed relationship for the other clinical responses.

**Table S6.** **Hospitalisation fatality risk (%) stratified by age and shielding groups during 1 January 2020 to 1 December 2020.** Midpoint 6 rounding was applied to numerator (number of deaths) and denominator (number of hospital admissions). The number of people in each age group is given in Table S1.

|  | Whole period | | Up to 30/06/2020 | | From 01/07/2020 | |
| --- | --- | --- | --- | --- | --- | --- |
| Age group  (yrs) | Non  shielding | Shielding | Non  shielding | Shielding | Non  shielding | Shielding |
| 0 - 4 | 0.0 | 0.0 | 0.0 | 0.0 | 0.0 | 0.0 |
| 5 - 11 | 1.5 | 0.0 | 0.0 | 0.0 | 2.7 | 0.0 |
| 12 - 17 | 0.9 | 0.0 | 2.6 | 0.0 | 0.0 | 0.0 |
| 18 - 29 | 0.5 | 1.6 | 0.4 | 3.0 | 0.3 | 3.4 |
| 30 - 39 | 1.6 | 2.9 | 2.5 | 1.9 | 1.0 | 2.0 |
| 40 - 49 | 4.0 | 9.6 | 5.0 | 10.4 | 2.7 | 7.4 |
| 50 - 59 | 8.4 | 13.6 | 12.2 | 14.8 | 3.9 | 12.6 |
| 60 - 69 | 18.2 | 22.1 | 24.9 | 25.7 | 10.8 | 18.1 |
| 70+ | 40.9 | 41.7 | 49.8 | 45.0 | 29.2 | 37.5 |

21 parameters (Table S5) were estimated jointly and their posterior medians and 95% CrI are reported (Table S7). We also include the Maximum a priori estimate (MAP) to give fuller information on the posterior distribution. These parameters included all epidemiological parameters of the model, 12 parameters for the age and shielding-status specific clinical responses *y*, *h*, and *d*, and two nuisance parameters (overdispersion of hospitalisation events and of death events). As the model parameters were correlated and as the temporal variation in the social contact data imposed strict constraints on which parameter values could capture the observed epidemic pattern, it was important to sample all parameters jointly rather than fixing some using literature values. This approach made sure as many data-compatible parameter value combinations as possible contributed to the posterior sample.

Convergence of each MCMC sampling chain was confirmed by inspection of the parameters posterior marginal densities (Figure S7) and traces (Figure S8), and by their Gelman-Rubin PSRF, which ranged between 1.002 and 1.061 across parameters, below the accepted upper cutoff of 1.1 (Gelman 2013). The effective sample size over all chains was 7455.

Regarding the estimated epidemiological parameters, the latent period posterior median of 2.9 days (Table S7) appears consistent with studies reporting a significant probability of PCR detection 1-3 days after infection ^12^. This parameter’s posterior median and 95% CrI are also consistent with the gamma distributions (with means 3.0 and 3.4 days) assumed in related model-fitting studies ^5,9^ based on information from literature sources. For the clinical infection period, our estimated posterior median of 3.8 days and 95% CrI (Table S7) are consistent with the mean infectious period of 4 days assumed in [23] and mean clinical infectious period of 3.8 days assumed in ^5^ based on information from independent literature sources. For the sub-clinical infection period, we estimated a median of 1.5 days and 95% CrI of [1.13,2.02], somewhat shorter than but consistent with the asymptomatic infection period mean of 2.9 days and 95% CrI [0.1, 10] assumed in ^5^. Our estimated *R_0_* posterior median of 2.2 and 95% CrI of [1.69, 2.80] (Table S7) are consistent with the reported means of 2.7 and 2.8 ^5,9^; and also with the reported mean of 1.95 and 95% CI [1.70, 2.26] in ^30^, estimated based not on the NGM, but on the Lotka-Euler equation and transmission pairs data from China in January-February 2020. Finally, for the infectiousness of unascertained infections in relation to clinical infection, *f*, we estimated a posterior median and 95% CrI of 0.186 (0.126, 0.292), which is close to the 0.223 assumed in ^31^ but much lower than the 0.5 assumed in ^32^. This result means that clinically infected people can be on average (95% CrI) 5.4 (3.4, 7.9) times more infectious than other infected people.

Estimates of other clinical progression parameters (Table S7), for the average periods from hospitalisation to recovery (*t_HR_*) and from hospitalisation to death (*t_HD_*), are in reasonable agreement with estimates in ^5^; our estimated average period from clinical infection to death in the community (*t_ID_*), median 14.9 days and 95% CrI [11.77, 18.43], is shorter than the assumed 22 days in ^4,5^.

Several factors in our estimation differed the from previous studies. For example, there is also difference in the fitted datasets of severe outcomes in England, and difference in the social contact data input to the models. In addition, we estimated all model parameters jointly without inputting literature values and used these to check the consistency of our estimates.

**Table S7. Estimated parameters of the transmission model.** Notation: Parameters that depend on age and shielding status have indices *i* and *s*. Each average duration or period is the inverse of the corresponding transition rate, e.g. *t_EI_* =1/*r_EI_*, where *r_EI_* is used directly in the model. Parameter *t_ID_* was estimated as the sum of two other sampled parameters (Table S5). MAP: Maximum-a-priori estimate. 95% CrI: 95% credible interval. The estimated parameter *p_I0_* is multiplied by the population size N to give the estimated number of clinically infected people at the start of the epidemic.

| **Symbol** | **Description** (each period is a mean duration) | **Median (95% CrI),** | **MAP** |
| --- | --- | --- | --- |
| *t_EI_, t_EU_* | Latent period | 2.86 (1.77,4.38) | 2.83 |
| *t_IR_* | Infectious period of clinical infections | 3.83 (3.36,4.34) | 3.58 |
| *t_UR_* | Infectious period of sub-clinical infections | 1.46 (1.13,2.02) | 1.66 |
| *R_0_* | Basic reproduction number | 2.18 (1.69,2.80) | 2.16 |
| *N_I0_ =*  *p_I0_ N* | Number of infected people on 27/01/2020. *N*=56.43 million | 148 (87,244), | 110 |
| *t_IH_, t_IO_* | Period to hospital admission from onset of clinical infection | Assumed equal to *t_IR_* |  |
| *t_HR_* | Period to recovery in hospital, from admission to discharge | mean (median) |  |
|  | Overall | 12.0 (6) |  |
|  | Up to 30 June 2020 | 10.1 (6) |  |
|  | From 1 July 2020 | 14.2 (7) |  |
| *t_HD_* | Period to mortality in hospital, from admission to death | mean (median) |  |
|  | Overall | 13.9 (10) |  |
|  | Up to 30 June 2020 | 12.5 (9) |  |
|  | From 1 July 2020 | 16.8 (12) |  |
| *t_ID_* | Period to mortality outside hospital from onset of critical infection | 14.87 (11.77,18.43), | 14.31 (*t_IO_* + *t_OD_*) |
| *t_OD_* | Period to mortality outside hospital from needing hospital admission | 11.02 (7.87,14.63), | 10,74 |
| *f* | Infectiousness of unascertained infection in relation to clinical infections | 0.186 (0.126, 0.292) | 0.171 |
| *β* | Probability of infection on contact | 0.71 (0.51,0.94), | 0.73 |
| y*_i.s_* | Probability of clinical symptoms in an infection (clinical fraction) | Figure S10, includes estimated amplitude and rate parameters for age groups 3 to 9 |  |
| h*_i.s_* | Probability of hospital admission while clinically infected | Figure S10, includes estimated amplitude and rate parameters |  |
| m*_i.s_* | Probability of death while in hospital, HFR | Table 2 |  |
| d*_i.s_* | Probability of death outside hospital while clinically infected | Figure S10, includes estimated amplitude and rate parameters |  |
| k*_H_* | Overdispersion of hospitalisation events | 3,07 (2.60,3.62) | 2.93 |
| k*_D_* | Overdispersion of death events | 1.02 (0.85,1.21) | 1.03 |

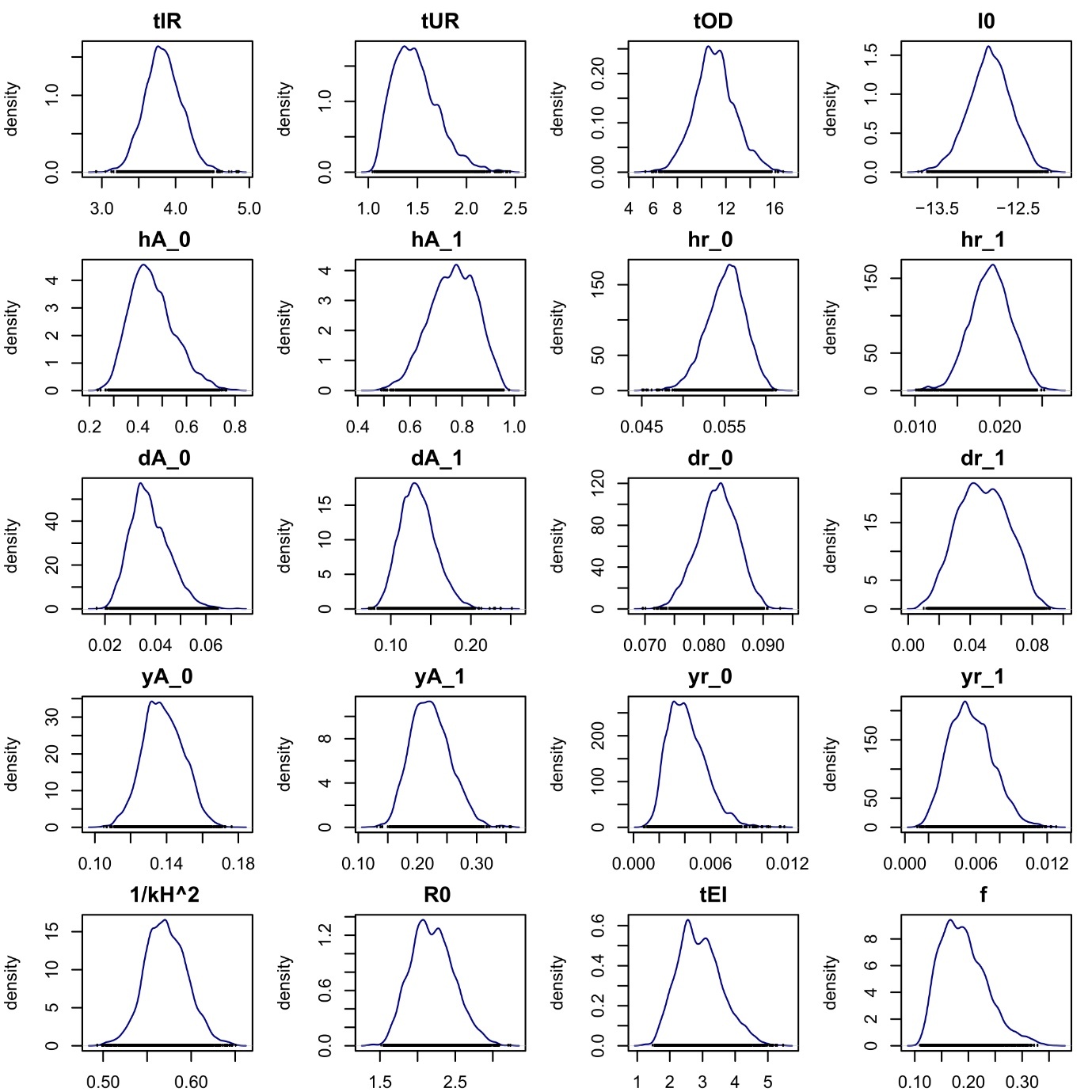
**Figure S7: Marginal posterior and prior distributions of parameters estimated using Bayesian MCMC sampling.** Parameters are described in Table S5 and ‘Model parameters’. The priors are Beta distributions described in Sec. 8, ‘Bayesian fitting’. Parameter *p_E0_* was log transformed. Subscripts ‘0’ and ‘1’ stand for ‘non-shielding’ and ‘shielding’, respectively. For clarity, the 21^st^ parameter (nuisance aggregation *k_D_*) is not shown.

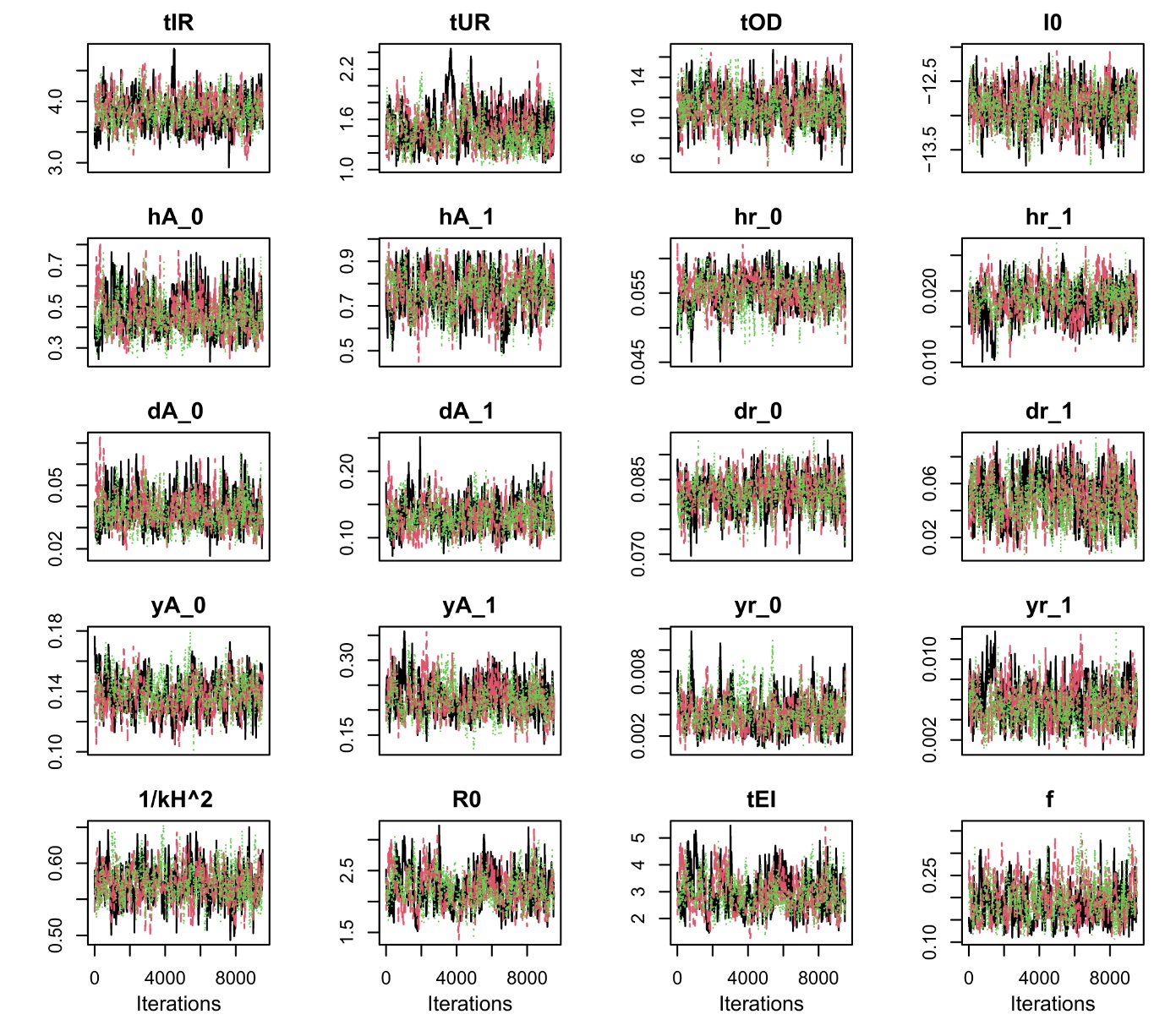
**Figure S8. Trace of parameters estimated using Bayesian MCMC sampling.** Parameters are described in Table S5 and ‘Model parameters’. Each trace shown has 3 sampling chains with over 9500 steps each. The traces are based on running each chain over 67,000 steps, discarding the first half and applying 1/7 thinning (Sec. 8, ‘Bayesian fitting’). Subscripts ‘0’ and ‘1’ stand for ‘non-shielding’ and ‘shielding’, respectively. For clarity, the 21^st^ parameter (nuisance aggregation *k_D_*) is not shown.

#### 11. Weekly incidence of severe COVID-19 outcomes during the pandemic

Further to discussion of Figure 2 in the main text, one marginal case of the fit regards deaths in hospital during wave two of the pandemic, for which the data are at the upper edge of the 95% PI (Figure 2 c-d). We expect wave two to be less well fitted by the model because several factors changed since wave one; while change in social contacts was captured by the CoMix survey data ^10^, change in other factors such as improved clinical outcomes ^3^ was only partially accounted for in the probability of death in hospital, *m* (Table 2), and in the rates of outcomes in hospital (Table S4). Another marginal case regards deaths outside hospital (Figure 2 e), where the data for the non-shielding population are relatively close to the upper 95% PI during wave one. Records of mortality outside hospital are likely to have some misreporting and to be less accurate about the cause of death, as testing would generally not have been applied or be applied less rigorously than in hospital. Therefore, we also expect these data to be less well fitted by the model.

Additionally to the discussion of Figure 3 in the main text, the apparent between-wave mismatch between the model median and data is an artifact of using two-period parameter values for the rates of recovery and death in hospital (Table S4 or S6). In addition, the two cases of marginal data inclusion in the model 95% PI seen in Figure 2 are also seen in Figure 3 for the same reasons as in Figure 2.

#### 12. Weekly incidence of severe outcomes by age and shielding status

The full, age-specific, model-predicted and observed, weekly incidence of severe COVID-19 outcomes in each shielding-status population (Figure S9), shows the data to which the model was fitted and their age-dependent temporal patterns. For clarity, only the model posterior median is shown without overlapping uncertainty intervals (Figure 3 displays uncertainty intervals for less refined age groupings that aggregate the age groups with very scarce events, see Sec. 8, ‘Bayesian fitting’).

These results show reasonable agreement between the fitted-model and the data across age groups; they also show consistency in how the model temporal patterns of incidence change across age groups. The following provisos apply: 1) the data are midpoint-6 rounded to prevent low-count disclosure (see Sec. 1), which shows in the younger age groups as truncated horizontal lines; 2) for age groups for which the weekly events recorded were too infrequent and irregular to inform the model, a fit to the total counts over the study period rather than to each time point was adopted (See Sec. 8).

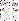

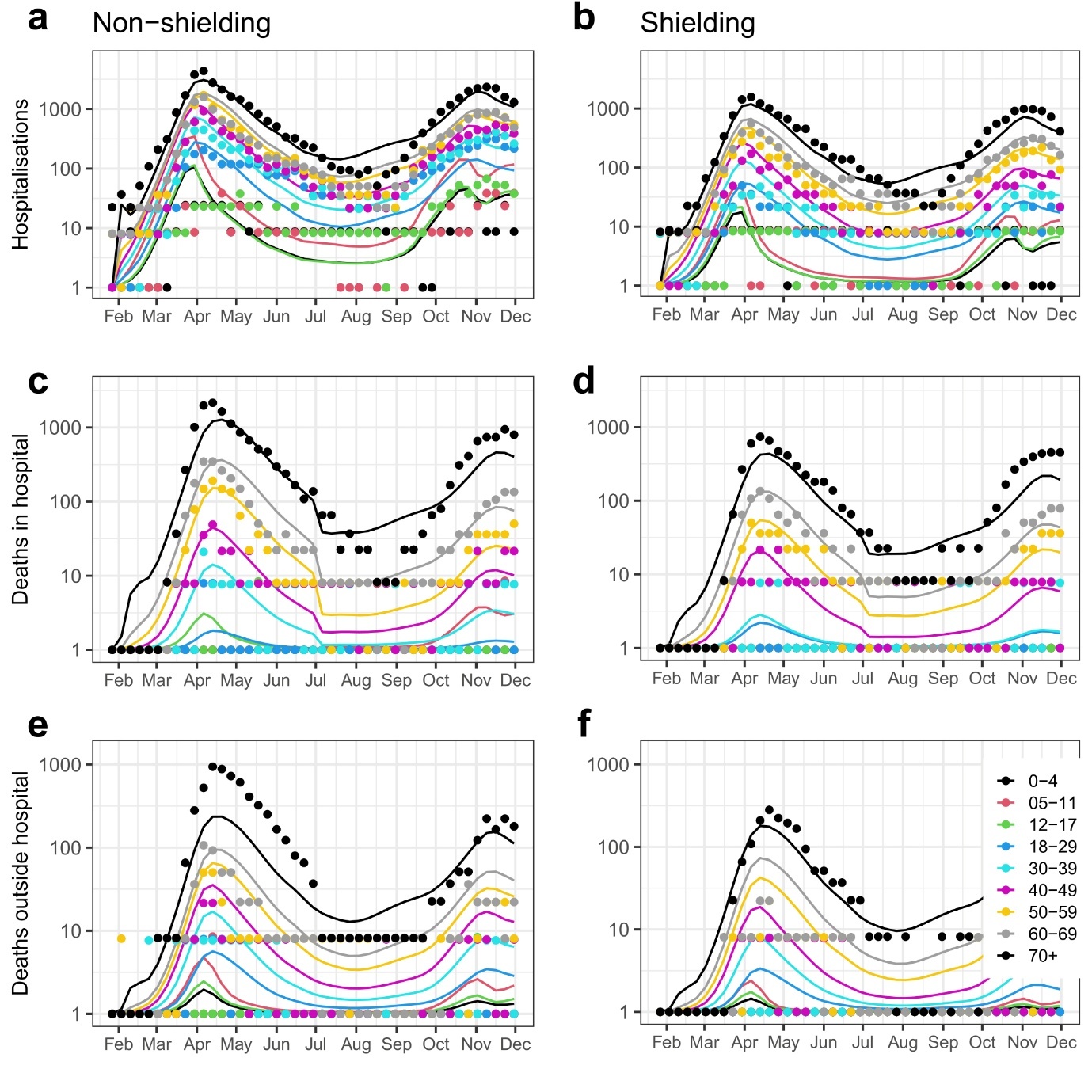

**Figure S9: National, age-specific weekly incidence of COVID-19 hospitalisations, deaths in hospital, and deaths outside hospital in England in the non-shielding and shielding populations**. The model and data are shown on a log-10 scale. Posterior predictive distribution of the fitted transmission model (median line) and patient EHR data (dots) from 27 January to 01 December 2020, for each age group and shielding status, aggregated over all NHS England regions. EHR data are scaled up from 24 million patients (general practices covered by the software provider TPP) to 56.6 million, the census population in England in mid 2020. The model was fitted to the age and shielding-status specific incidence of each clinical outcome; posterior predictive distributions were derived by drawing parameters from the joint posterior sample and simulating observations. Prediction intervals are not shown (unlike Figure 3) for clarity and due to low weekly counts (and their lack of temporal pattern) in younger age groups (Sec. 8, ‘Bayesian fitting’). For plotting on log10 scale, a lower cut-off of 1 was applied.

#### 13. Clinical responses to COVID-19 by age and shielding status

The probabilities of developing clinical outcomes from COVID-19 (Table S2), i.e. developing clinical symptoms, being hospitalised, or dying, by age and by shielding status, were used in the transmission model to calculate the rates at which infected people acquired severe outcomes from COVID-19 (See Sec. 3, ‘Transmission model’).

Further to discussion in the main text, Figure S10 shows that the probabilities of hospitalisation if clinically infected were much higher in shielding people, and the probabilities of developing clinical symptoms were also higher in shielding people (Figure S10a-d). However, the probability of an infection becoming clinical and the probability of hospitalisation of a clinically symptomatic person are negatively correlated and, therefore, it is statistically difficult to identify both parameters jointly. Therefore, we also plotted the probability of an infection being clinical and requiring hospitalisation, i.e. the infection hospitalisation risk or IHR, given by the product of the latter probabilities, which we expected to be more directly conditioned by the data and to be more directly interpretable. This probability was also higher for shielding people (Figure S10e-f). These results are consistent with the expectation that shielding people were at greater clinical risk from COVID-19. Furthermore, the probability of death in hospital (Figure S10i-j) did not differ notably between shielding and non-shielding people up to June 2020, but was higher for shielding people after 1 July 2020. The probability of dying outside hospital was considerably higher for shielding people (Figure S10g-h). Some of the probabilities in Figure S10 were estimated in other model-based studies ^5,9^ for the general population (not shielding stratified).

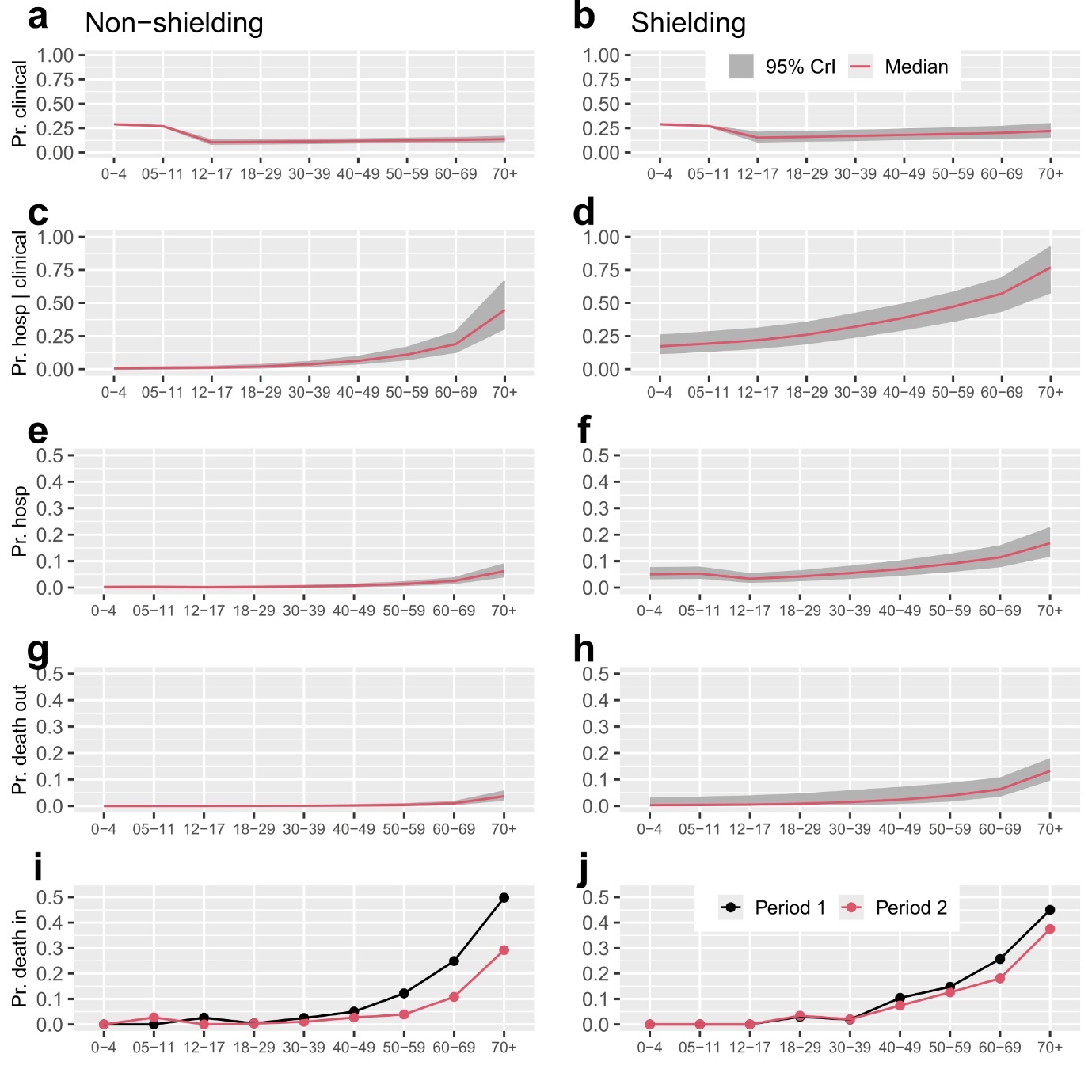

**Figure S10: Clinical response probabilities in COVID-19-affected patients by age and shielding status.** Marginal posterior distributions of the probabilities (median and 95% credible interval (95% CrI)), and EHR-based proportions (last row) from 27 January to 30 June 2020 (Period 1) and from 01 July to 01 December 2020 (Period 2). a-b: Probability of an infection causing clinical symptoms (*y_i,s_*, Pr. clinical). c-d: Probability of hospitalisation of a person with clinical symptoms (*h_i,s_*, Pr. hosp | clinical). e-f: Probability of being clinically infected and hospitalised (*y_i,s_ x h_i,s_*, Pr. hosp), i.e. the infection hospitalisation risk. g-h: Probability of death outside hospital (*d_i,s_*, Pr. death out). i-j: Probability of death in hospital (*m_i,s_*, Pr. death in). In a-b, the values of the curve for the first two age groups are from literature [16] and do not change with shielding status, unlike those for age groups 3 to 9 (see Sec. 4, 'Model parameters').

#### 14. Basic reproductive number over time

During the study period, from 27 January to 01 December 2020, there is temporal variation in the basic reproduction number, *R_0_* (Figure S11). This variation is due to temporal variation in the social contacts among age groups (Figure 1). *R_0_* in week 1 is the estimated marginal posterior distribution (Table S7). In subsequent weeks, *R_0_* varied because social contacts varied. Reasons for calling this time-varying parameter *R_0_* rather than *R_t_* are discussed in Sec. 10, ‘Estimated model parameters and MCMC diagnostics’.

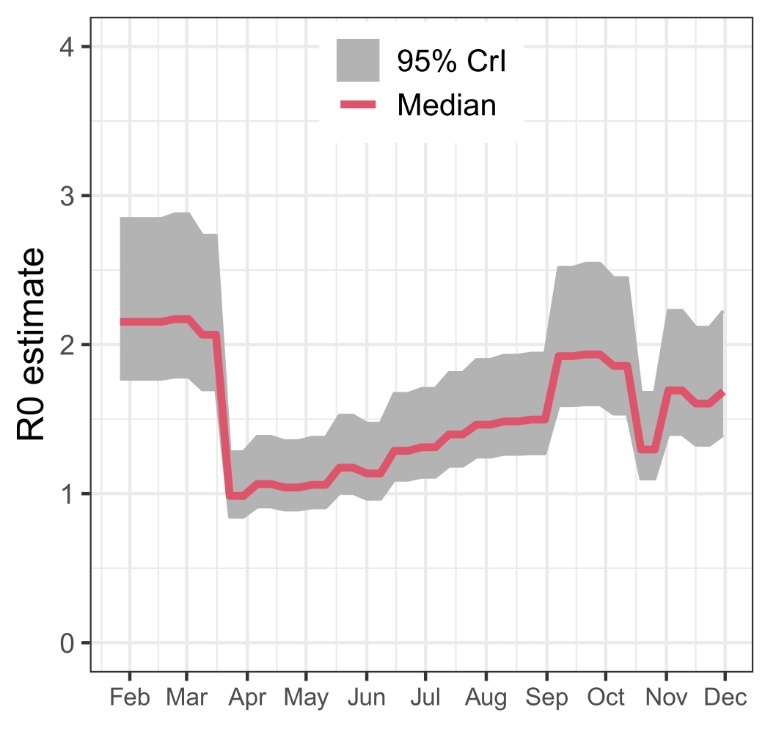

**Figure S11: Basic reproductive number over time.** Posterior median and 95% credible interval (95% CrI), from fitting the transmission model, over the period of 27 January to 01 December 2020 (Sec. 7, ‘Basic reproduction number’).

#### 15. Sensitivity Analysis 1: Poisson vs Negative Binomial contacts

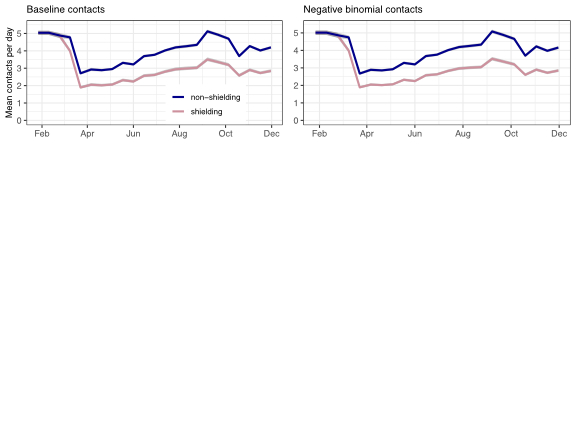

**Figure S12: Number of contacts through time in the baseline model (Poisson) and when using a Negative Binomial distribution.** Note model is fitted to age-specific values, but they are aggregated here for clarity.

**Table S8. Parameter estimates from the main model and that where contacts were assumed to follow a Negative Binomial distribution.** Median and 95% CIs are shown. *t_ID_* and *β* are derived parameters.

|  | **Main Model** |  | **Negative Binomial model** | |
| --- | --- | --- | --- | --- |
| Parameter | Median | 95% CI | Median | 95% CI |
| f | 0.186 | [0.126, 0.292] | 0.179 | [0.121, 0.347] |
| R0 | 2.2 | [1.687, 2.799] | 2.1 | [1.663, 2.672] |
| tEI | 2.9 | [1.767, 4.378] | 2.6 | [1.686, 3.797] |
| tUR | 1.5 | [1.134, 2.017] | 1.4 | [1.118, 1.993] |
| tIR | 3.8 | [3.359, 4.335] | 4.0 | [3.512, 4.693] |
| tIH | 3.8 | [3.359, 4.335] | 4.0 | [3.512, 4.693] |
| tOD | 11.0 | [7.871,14.629] | 11.0 | [7.799, 14.378] |
| (tID derived) | 14.9 | [11.771,18.427] | 15.0 | [11.856, 18.257] |
| (*β* derived) | 0.729 | [0.514, 0.944] | 0.669 | [0.488, 0.926] |
| kH | 3.1 | [2.599, 3.622] | 3.1 | [3.574, 2.617] |
| kD | 1.0 | [0.845, 1.208] | 1.0 | [1.199, 0.845] |
| I0 | 148 | [87, 244] | 151 | [87, 250] |

**Table S9. Outcomes averted in the main model and that where contacts were assumed to follow a Negative Binomial distribution.** Median and 95% CrI are given.

|  |  | **Hospitalisations** | | **Deaths** |  |
| --- | --- | --- | --- | --- | --- |
|  |  | **Shielding** | **Non-Shielding** | **Shielding** | **Non-Shielding** |
| **Main Model** | **Count** | 9100  [7800, 10600] | 17700  [14700 ,21800] | 2800  [2300, 3500] | 4500  [3700, 5500] |
|  | **Percent** | 25% [24,28] | 13% [12,16] | 23% [21,25] | 13% [11,15] |
| **Negative Binomial model** | **Count** | 9200  [8000, 10800] | 18000  [14900, 22000] | 2900  [2400, 3600] | 4600  [3800, 5600] |
|  | **Percent** | 26% [24,28] | 14% [12,16] | 23% [21,25] | 13% [12,15] |

#### 16. Sensitivity analysis 2: Further reduction in contacts

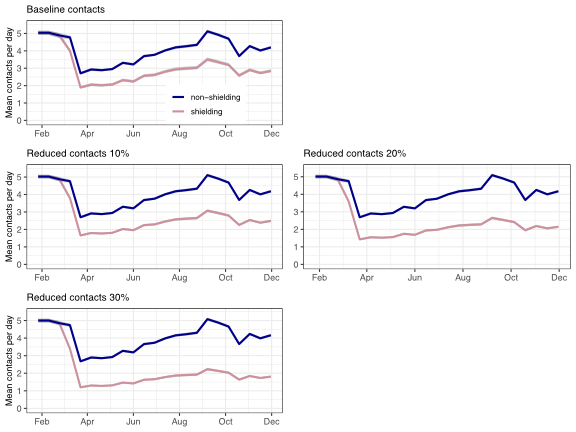

**Figure S13: Number of contacts through time in the baseline model and when assuming contacts are reduced by a further 10% or 20% compared to the CoMix study.** Note model is fitted to age-specific values, but they are aggregated here for clarity.

**Table S10. Parameter estimates from the main model and that when assuming contacts are reduced by a further 10% or 20% compared to the CoMix study.** Median and 95% CIs are shown. *t_ID_* and *β* are derived parameters.

|  | Main Model | | 10% Further Reduction | | 20% Further Reduction | |
| --- | --- | --- | --- | --- | --- | --- |
| Parameter | Median | 95% CI | Median | 95% CI | Median | 95% CI |
| f | 0.186 | [0.126, 0.292] | 0.192 | [0.124, 0.325] | 0.195 | [0.129, 0.3] |
| R0 | 2.2 | [1.687, 2.799] | 2.2 | [1.672, 2.7922] | 2.2 | [1.758, 2.888] |
| tEI | 2.9 | [1.767, 4.378] | 2.8 | [1.709, 4.3097] | 3.0 | [1.892, 4.566] |
| tUR | 1.5 | [1.134, 2.017] | 1.5 | [1.141, 1.966] | 1.5 | [1.134, 1.955] |
| tIR | 3.8 | [3.359, 4.335] | 3.9 | [3.438, 4.5313] | 3.9 | [3.409, 4.506] |
| tIH | 3.8 | [3.359, 4.335] | 3.9 | [3.438, 4.5313] | 3.9 | [3.409, 4.506] |
| tOD | 11.0 | [7.871,14.629] | 11.0 | [7.493, 14.418] | 11.0 | [7.767,14.226] |
| (tID derived) | 14.9 | [11.771,18.427] | 14.9 | [11.746,18.199] | 15.0 | [11.892,18.035] |
| (*β* derived) | 0.729 | [0.514, 0.944] | 0.681 | [0.486, 0.954] | 0.698 | [0.504, 0.96] |
| kH | 3.1 | [2.599, 3.622] | 3.1 | [2.621, 3.634] | 3.1 | [2.614, 3.562] |
| kD | 1.0 | [0.845, 1.208] | 1.0 | [0.855, 1.201] | 1.0 | [0.833, 1.194] |
| I0 | 148 | [87, 244] | 143 | [80, 237] | 134 | [77, 224] |

**Table S11. Outcomes averted in the main model and that where contacts were reduced by 10, 20 and 30% beyond the CoMix study.** Median and 95% CrI are given.

|  |  | **Hospitalisations** | | **Deaths** |  |
| --- | --- | --- | --- | --- | --- |
|  |  | **Shielding** | **Non-Shielding** | **Shielding** | **Non-Shielding** |
| **Main Model** | **Count** | 9100  [7800, 10600] | 17700  [14700, 21800] | 2800  [2300, 3500] | 4500  [3700, 5500] |
|  | **Percent** | 25% [24,28] | 13% [12,16] | 23% [21,25] | 13% [11,15] |
| **10% Further Reduction** | **Count** | 14600  [12700, 17200] | 27600  [22800, 35000] | 4700  [3900, 5900] | 7300  [6000, 6100] |
|  | **Percent** | 35% [33,38] | 19% [17,23] | 31% [31,36] | 20% [17,23] |
| **20% Further Reduction** | **Count** | 21400 [18500,25100] | 38700 [31800,48900] | 7047 [5900,8800] | 10500 [8700,13300] |
|  | **Percent** | 45% [42,48] | 25% [22,29] | 42% [40,46] | 26% [23,30] |

2	OpenCodelists. https://www.opencodelists.org/.

3	UK Government, Department of Health & Social Care. [treatment-improved] Technical report on the COVID-19 pandemic in the UK, Chapter 10: improvements in care of COVID-19. 2023 https://www.gov.uk/government/publications/technical-report-on-the-covid-19-pandemic-in-the-uk/chapter-10-improvements-in-care-of-covid-19 (accessed Dec 19, 2022).

4	Davies NG, Kucharski AJ, Eggo RM, *et al.* Effects of non-pharmaceutical interventions on COVID-19 cases, deaths, and demand for hospital services in the UK: a modelling study. *Lancet Public Health* 2020; **5**: e375–85.

5	Knock ES, Whittles LK, Lees JA, *et al.* Key epidemiological drivers and impact of interventions in the 2020 SARS-CoV-2 epidemic in England. *Sci Transl Med* 2021; **13**: eabg4262.

6	Keeling MJ, Dyson L, Guyver-Fletcher G, *et al.* Fitting to the UK COVID-19 outbreak, short-term forecasts and estimating the reproductive number. *Stat Methods Med Res* 2022; **31**: 1716–37.

7	Williamson EJ, Walker AJ, Bhaskaran K, *et al.* Factors associated with COVID-19-related death using OpenSAFELY. *Nature* 2020; **584**: 430–6.

8	Herrera-Esposito D, de los Campos G. Age-specific rate of severe and critical SARS-CoV-2 infections estimated with multi-country seroprevalence studies. *BMC Infect Dis* 2022; **22**: 311.

9	Davies NG, Klepac P, Liu Y, *et al.* Age-dependent effects in the transmission and control of COVID-19 epidemics. *Nat Med* 2020; **26**: 1205–11.

10	Gimma A, Munday JD, Wong KLM, *et al.* Changes in social contacts in England during the COVID-19 pandemic between March 2020 and March 2021 as measured by the CoMix survey: A repeated cross-sectional study. *PLOS Med* 2022; **19**: e1003907.

11	Davies NG, Barnard RC, Jarvis CI, *et al.* Association of tiered restrictions and a second lockdown with COVID-19 deaths and hospital admissions in England: a modelling study. *Lancet Infect Dis* 2021; **21**: 482–92.

12	Hellewell J, Russell TW, Matthews R, *et al.* Estimating the effectiveness of routine asymptomatic PCR testing at different frequencies for the detection of SARS-CoV-2 infections. *BMC Med* 2021; **19**: 106.

13	Birrell P, Blake J, Van Leeuwen E, Gent N, De Angelis D. Real-time nowcasting and forecasting of COVID-19 dynamics in England: the first wave. *Philos Trans R Soc B Biol Sci* 2021; **376**: 20200279.

14	Google Mobility. https://www.google.com/covid19/mobility/ (accessed Nov 24, 2025).

15	https://www.visualcrossing.com/. https://www.visualcrossing.com/. https://www.visualcrossing.com/.

16	Munday, James D, Jarvis, Christopher I, Gimma, Amy, *et al.* [CoMix-zenodo] CoMix - Age structured contact matrices for 9 key periods of the COVID-19 epidemic in England (Version v1). 2021; published online April 9. https://doi.org/10.5281/zenodo.4677018 (accessed Dec 3, 2024).

17	National Audit Office. [shielding-start] Protecting and supporting the clinically extremely vulnerable during lockdown. 2021 https://www.nao.org.uk/reports/protecting-and-supporting-the-vulnerable-during-lockdown/ (accessed Jan 24, 2025).

18	UK Government, Ministry of Housing, Communities and Local Government. [shielding-start2] Shielding clinically vulnerable people from COVID-19. 2020 https://www.local.gov.uk/sites/default/files/documents/SHIELDING%20GUIDANCE%20AND%20FAQS%20COMBINED%20-%2024%20APRIL%202020.pdf (accessed Jan 24, 2025).

19	Sullivan O, Gershuny J. UKTUS; UK Time Diary StudyUnited Kingdom Time Use SurveyUnited Kingdom Time Use Survey, 2014-2015. 2023. DOI:10.5255/UKDA-SN-8128-1.

20	Diekmann O, Heesterbeek JAP, Metz JAJ. On the definition and the computation of the basic reproduction ratio R0 in models for infectious diseases in heterogeneous populations. *J Math Biol* 1990; **28**: 365–82.

21	Braak, Caro J. F. ter. A Markov Chain Monte Carlo version of the genetic algorithm Differential Evolution: easy Bayesian computing for real parameter spaces. *Stat Comput* 2006; **16**: 239–49.

22	Brooks, S, Gelman, A, Jones, G. (eds). Handbook of Markov Chain Monte Carlo. New York: Chapman and Hall/CRC, 2011.

23	Braak CJF ter, Vrugt JA. Differential Evolution Markov Chain with snooker updater and fewer chains. *Stat Comput* 2008; **18**: 435–46.

24	Hartig, F., Minunno, F., Paul, S., Cameron, D., Ott, T., Pichler, M. BayesianTools: General-Purpose MCMC and SMC Samplers and Tools for Bayesian Statistics. . . 2023; **version 0.1.8**. DOI:10.32614/CRAN.package.BayesianTools.

25	De Angelis D, Presanis AM, Birrell PJ, Tomba GS, House T. Four key challenges in infectious disease modelling using data from multiple sources. *Chall Model Infect Dis Dyn* 2015; **10**: 83–7.

26	Andrews, Colm D., Schultze A, Curtis H, *et al.* OpenSAFELY: Representativeness of electronic health record platform OpenSAFELY-TPP data compared to the population of England. *Wellcome Open Res* 2022; **7**: 191.

27	Gelman, A., Carlin, J.B., Stern, H.S. Bayesian data analysis, Third. New York: Chapman and Hall/CRC, 2013.

28	Office for Nationa Statistics. [ONS-CIS-data] Coronavirus (COVID-19) Infection Survey: England. 2023; published online March 10. https://www.ons.gov.uk/peoplepopulationandcommunity/healthandsocialcare/conditionsanddiseases/datasets/coronaviruscovid19infectionsurveydata (accessed March 14, 2024).

29	Office for Nationa Statistics. [ONS-CIS-methods] Coronavirus (COVID-19) Infection Survey: methods and further information. 2023 https://www.ons.gov.uk/peoplepopulationandcommunity/healthandsocialcare/conditionsanddiseases/methodologies/covid19infectionsurveypilotmethodsandfurtherinformation (accessed March 14, 2024).

30	Chen C-J, Lee P-I, Chang S-C, *et al.* Seroprevalence and severity of 2009 pandemic influenza A H1N1 in Taiwan. *PloS One* 2011; **6**: e24440.

31	Knock ES, Whittles LK, Lees JA, *et al.* Key epidemiological drivers and impact of interventions in the 2020 SARS-CoV-2 epidemic in England. *Sci Transl Med* 2021; **13**: eabg4262.

32	Davies NG, Klepac P, Liu Y, *et al.* Age-dependent effects in the transmission and control of COVID-19 epidemics. *Nat Med* 2020; **26**: 1205–11.
